# Self- and Caregiver-Reported Choice-Making in Autistic Adults: Development and Validation of the AASPIRE–Choices and Decisions Scale

**DOI:** 10.64898/2026.05.07.26352693

**Authors:** So Yoon Kim, Kristen Gillespie-Lynch, Steven K. Kapp, Liu-Qin Yang, Anna “Furra” Wallington, Dora M. Raymaker, Ian Moura, Katherine McDonald, Joelle Maslak, Rachel Kripke-Ludwig, Andrea Joyce, Willi Horner-Johnson, Emanuel Frowner, Mary Baker-Ericzén, Christina Nicolaidis

**Author notes:** **Corresponding author:** Christina Nicolaidis, School of Social Work, Portland State University, Portland, OR 97201, USA.

## Abstract

Self-determination has been assessed as an internal psychological construct. External factors may also affect self-determination, but opportunities to make choices and decisions remain understudied. We developed and evaluated the AASPIRE-Choices and Decisions Scale (AASPIRE-CDS), a new measure of autistic adults’ opportunities to make choices and decisions, using a community-based participatory approach. We created and refined the AASPIRE-CDS through an iterative process. Data, from the AASPIRE Outcomes Project, included 839 autistic adults participating through direct report, supported direct report, and caregiver report (CR). Exploratory and confirmatory analyses supported a unidimensional structure. Measurement invariance analyses supported configural, metric, and partial scalar invariance across report type without CR, and across living status, with and without CR. The AASPIRE-CDS showed high internal consistency, test-retest reliability, and preliminary responsiveness to change over time. Convergent validity analyses showed that higher AASPIRE-CDS scores were associated with greater self-determination and communication fluency, more independent living, and fewer support needs. The AASPIRE-CDS showed weaker (albeit significant) associations with quality of life, overall health, and employment satisfaction than the self-determination measure showed with these variables. This pattern suggests that opportunities for choice-making are related to, but distinct from, commonly used measures of self-determination. Findings support the AASPIRE-CDS as a valid and reliable measure of choice-making opportunities in autistic adults across support needs but suggest caution interpreting CR. They underscore the importance of supporting autistic adults’ choice-making and evaluating opportunities for choice alongside internal self-determination. Future research should use larger CR samples to examine the validity of caregiver-reported choice-making opportunities.

**Lay Summary:** Self-determination means having control over your own life and being able to make choices. We created a new measure to better understand whether autistic adults have real opportunities to make their own choices in everyday life. Findings show the importance of giving autistic adults the support they need to make choices and making sure they have real opportunities to make choices.

## Introduction

Self-determination means “acting as the primary causal agent in one’s life and making choices and decisions regarding one’s quality of life (QoL) free from undue external influence” (Wehmeyer et al., 1996, p. 24). Greater self-determination is associated with increased well-being and QoL among autistic and non-autistic people (Kim, 2019; Wehmeyer, 2021). Autism research frequently conceptualizes self-determination as a psychological construct centered on motivational processes involved in decision-making (Morán et al., 2021). Such approaches often emphasize individual-level difficulties, such as differences in executive functioning, that may interfere with autistic adults’ goal-setting, planning, and follow-through (Thompson-Hodgetts et al., 2023).

Consistent with this framing, some studies have reported that autistic people may report lower self-determination than peers without disabilities or with other developmental disabilities when self-determination is measured as an individual psychological construct (Chou et al., 2017; Shogren et al., 2018). However, this pattern may partly reflect misunderstandings or constraints imposed by others, including societal assumptions that autistic people have reduced cognitive agency and capacity for planning and self-directed action (e.g., self-control; Kim et al., 2025), which may in turn limit their opportunities to exercise self-determined behavior.

### Reframing Self-Determination as Shaped by Choice and Decision-Making Opportunities

Because self-determined behavior must occur free from undue external control (Wehmeyer et al., 1996), access to meaningful decision-making opportunities is essential. Indeed, Self-Determination Theory (Ryan & Deci, 2002) identifies autonomy, competence, and relatedness as fundamental psychological needs essential for self-determination and well-being. Here, we conceptualize autonomy as acting (or choosing not to act) in accord with one’s authentic interests, values, and desires, rather than as independence, as autonomous choices are often supported by other people (Chirkov et al., 2003). Opportunities to make autonomous choices and decisions, that is, having chances to select among alternatives and determine courses of action, ranging from everyday choices, such as what to eat, to major life decisions, such as what career to pursue, have long been recognized as key indicators of well-being and social inclusion among people with disabilities (Kishi et al., 1988), including autism (Ryan et al., 2024). This includes the opportunity to make potentially harmful choices and decisions (i.e., the dignity of risk: Mukherjee, 2022).

However, prior studies have reported that autistic people tend to make fewer choices than people with other disabilities (Mehling & Tassé, 2015; Song et al., 2026). These reduced opportunities may be shaped not only by individual characteristics but also by interpersonal and structural conditions. In some cases, parents pursue guardianship for autistic individuals with high support needs, highlighting the complex and often emotional balancing act between protection and autonomy (Ferguson et al., 2025). While parental advocacy can play a vital role in promoting self-determination for autistic youth with and without intellectual disability (Moser et al., 2025), some autistic youth report that even well-meaning parental involvement can feel overprotective (Nachman, 2024). Together, these examples underscore the importance of examining opportunities for choice and decision-making among autistic people, as these opportunities may not be fully captured by approaches that frame self-determination primarily as an internal psychological construct.

### Instruments Measuring Choices and Decisions

Several instruments, many of which were initially developed for people with intellectual disability, have been validated and used to evaluate self-determination in autistic people, including the Self-Determination Inventory: Adult Report (SDI:AR; Shogren et al., 2021) and the ARC’s Self-Determination scale (Stancliffe, 2020; Wehmeyer, 1995). However, existing self-determination measures do not primarily focus on opportunities to make choices and decisions.

Existing instruments focusing on choice-making opportunities were developed and validated for people with intellectual and/or developmental disabilities rather than for autistic people. Factor analyses of these choice measures often reveal a distinction between everyday choices and major life choices; people with intellectual disability typically have less access to the latter than the former (Stancliffe, 2020). However, these instruments also have limitations. They include a similar set of items because they were developed by adapting earlier instruments that focused on common everyday choices that apply to most respondents and therefore minimize missing data (Stancliffe, 2020). Some particularly significant personal decisions, e.g., about sexuality, are missing entirely from most choice scales, yet such choices are highly consequential and constitute a key human right (United Nations, 2006).

Some surveys designed to assess disability service systems’ performance and quality include scales intended to evaluate the degree to which service recipients are supported in making choices and decisions. Most notably, the U.S.-based National Core Indicators–Intellectual and Developmental Disabilities (NCI-IDD) system, an optional state service-outcomes monitoring system for people receiving publicly funded developmental disability services and administered using standardized protocols and trained interviewers, includes a Choices and Decision-Making subdomain with two intended subscales: Life Decisions and Everyday Choices (Human Services Research Institute & National Association of State Directors of Developmental Disabilities Services [HSRI & NASDDDS], 2025). Although developers report strong reliability and validity for the overall NCI-IDD, the survey was designed primarily to evaluate and improve system-level performance in publicly funded developmental disability services rather than to assess individual, self-reported outcomes (HSRI & NASDDDS, 2025).

Researchers sometimes conduct secondary analyses of current and earlier versions of NCI-IDD data, yet this body of research also highlights the need for a valid measure of choice and decision-making. For example, exploratory factor analysis (EFA) of national NCI data from 2009 to 2011 found limited evidence for a hypothesized “choices” dimension among autistic adults and adults with developmental disabilities other than autism (Mehling & Tassé, 2015). Instead, choice-related items were distributed across two dimensions: social determination, reflecting choice in everyday activities such as how to spend free time, and personal control, reflecting choice over support arrangements such as who provides help at home. However, when Jones and colleagues (2018) administered the NCI alongside the short form of the ARC’s Self-Determination Scale to individuals with intellectual and developmental disabilities in Oklahoma, they found no evidence for the personal control dimension documented by Mehling and Tassé. Instead, they identified a dimension reflecting opportunities for autonomy across a broad range of everyday, service-related, and participatory domains, which was associated with higher self-determination. These findings suggest that the factor structure of choice-related opportunities in the NCI may not be reliable. Jones and colleagues (2018) also emphasized the need for future research on the comparability of proxy and self-report data, particularly because participants who did not report at least two choices on their own were excluded from their study due to concerns about the reliability of proxy reports.

In sum, there is a need for a measure of choice and decision-making for autistic people that is designed to assess this construct as an individual outcome rather than an indicator of service quality or system performance; that is intended for individuals beyond publicly funded developmental disability services; that can be used in research without intensive interviewer training or highly structured administration procedures; that shows a consistent structure across studies; and for which evidence on the comparability of self-report and proxy report is available.

### The Current Study

As a part of the AASPIRE Outcomes Project, a Community-Based Participatory Research (CBPR) initiative that brings together autistic and non-autistic academics and community members to identify outcomes that matter most to autistic adults, develop adapted and de novo community-supported outcome measures, and inform improvements in healthcare and disability services for autistic adults (Nicolaidis et al., 2020, 2025), this study focuses on the development and psychometric properties of the newly developed AASPIRE Choices and Decisions Scale (AASPIRE-CDS). We describe the minor adaptations to the Self-Determination Inventory: Adult Report-AASPIRE (SDI:AR-AASPIRE) and its psychometric findings in Supplementary Materials A, including a list of changes made from the SDI:AR, its factor structure, measurement invariance, latent mean and variance comparisons, and internal consistency. Because the AASPIRE-CDS is a newly developed measure, whereas the SDI:AR-AASPIRE is an adaptation of an existing instrument, the present article focuses primarily on the development and psychometric evaluation of the AASPIRE-CDS.

This study examines the content validity, factor structure, measurement invariance, construct validity, internal consistency, test-retest reliability, and responsiveness to change of the AASPIRE-CDS. We evaluated measurement invariance for AASPIRE–CDS across two variables that may influence how respondents interpret or use the response scale, report type and living status. Previous studies have shown no to moderate agreement between self- and proxy reports on autonomy- and choice-related measures (Shogren et al., 2021; Stancliffe & Parmenter, 1999; Tomaszewski et al., 2022). Given that self-reports reflect internal states such as preferences and intentions, whereas proxy reports rely on observable behavior or assumptions about capacity (Keith et al., 2019), we follow prior research (e.g., Lin et al., 2013; Shogren et al., 2021) by examining measurement invariance based on response type. Additionally, living situations may also shape how autistic adults interpret items and use response options. Autistic adults living independently generally encounter more frequent opportunities to make choices and decisions, whereas congregate and institutional settings often impose autonomy-limiting constraints (Kozma et al., 2009; Tichá et al., 2012; Wehmeyer & Metzler, 1995). In shared living contexts (e.g., living with parents or in residential settings), constraints on choice may be more common and sometimes unavoidable because decisions must take others’ needs and preferences into account. As a result, autistic adults in these contexts may rate situations involving partial or shared decision-making as reflecting relatively high choice, whereas those living independently might rate the same situations as reflecting less choice.

For convergent validity, we hypothesized that higher AASPIRE-CDS scores would be associated with more independent living contexts (Al Ansari et al., 2024) and greater communication fluency (Kapp, 2023) and fewer independent living support needs (Cheak-Zamora et al., 2022; Kaplan-Kahn et al., 2025) and personal care needs (Ferguson et al., 2025), as these indicators reflect external conditions and/or support needs that may shape the extent to which individuals are able to make choices. We expected the AASPIRE-CDS and SDI:AR-AASPIRE to be positively correlated, reflecting related but distinct constructs, but not so highly correlated as to indicate redundancy. Further, to examine discriminant validity, we tested whether correlations with subjective outcome indicators theoretically associated with self-determination (i.e., employment satisfaction, overall health, and QoL, Kim, 2019) were stronger for the SDI:AR-AASPIRE than for the AASPIRE-CDS.

## Methods

### Selection of Choice Construct and Instrument Development

Phase 1 of the AASPIRE Outcomes Project used a CBPR-nested Delphi process with experts with academic, professional, and lived experience to identify high-priority outcomes and evaluate candidate measures (Nicolaidis et al., 2025). Although self-determination was identified as a priority outcome, panelists indicated that existing instruments did not adequately capture what many considered most important: whether autistic adults had meaningful opportunities to make choices and decisions in everyday life. We therefore concluded that self-determination should be assessed using two complementary measures: an adapted version of the SDI:AR-AASPIRE to assess internal psychological aspects of self-determination and a separate measure to assess opportunities to make choices.

In Phase 2, a team of autistic and non-autistic partners collaboratively created high-priority outcome measures through an iterative CBPR process that included cognitive interviews with 17 autistic adults and six caregivers and iterative measure refinement (Nicolaidis et al., 2020). Initial feedback indicated that items organized around broad life domains with multiple examples were difficult to answer. We therefore revised the new instrument so that each item focused on a single specific choice or decision, with items ranging from lower-stakes everyday choices (e.g., what to wear) to higher-stakes decisions (e.g., where to live). Feedback also suggested that framing the construct as “freedom” to make choices could imply that greater choice is always preferable or appropriate. In response, we revised the wording to focus on whether a person “gets to” make a given choice or decision and added prefatory language acknowledging that people may not always be able or expected to make all decisions related to their own lives. Cognitive interviews with the revised measure were substantially more positive, supporting its content validity. We provide additional details regarding Phases 1 and 2 of our overall project in Supplementary Materials B, including the CBPR-nested Delphi process, review of existing choice measures, cognitive interview participants and their feedback, and resulting item revisions. The resulting instruments included direct-report (DR) and caregiver-report (CR) versions; in the CR version, caregivers served as informants, and we adapted the items by changing them to third-person phrasing and adding “I think” to the beginning of each item.

In Phase 3, we conducted a prospective cohort study of autistic adults who completed the de novo and adapted instruments, including the AASPIRE-CDS. This study used data collected in Phase 3 for the validation analyses.

### Participants

In the prospective cohort study, we recruited 870 participants from three subcohorts (Healthcare, Disability Services Systems, and Community; see Nicolaidis et al., 2026a). Participants completed a multi-instrument online survey that included measures of 22 outcomes identified as priorities through the AASPIRE Outcomes Project, as well as demographic and disability-related information. Autistic participants could complete the DR version independently (DR sample) or with assistance from a supporter (direct-report caregiver-supported [DR-CS] sample). When autistic participants or caregivers reported during screening that an autistic participant could not answer the survey directly, even with support, caregivers completed the CR version as informants (CR sample). Among the CR respondents, 63 completed the relationship item. Most identified as parents (*n* = 60), with smaller numbers identifying as a sibling (*n* = 1), friends (*n* = 2), direct support providers (*n* = 9), or other (*n* = 7; e.g., guardian). Respondents could endorse more than one category. We assigned report type at the overall survey level rather than separately for each instrument. Three participants classified as DR-CS indicated that someone answered the AASPIRE-CDS questions without their input. We repeated the analyses after excluding these participants and obtained results consistent with the primary analyses. After excluding 31 participants who had missing responses for all AASPIRE–CDS items, 839 participants are represented in the following analyses (see Table 1).

**Table 1.**
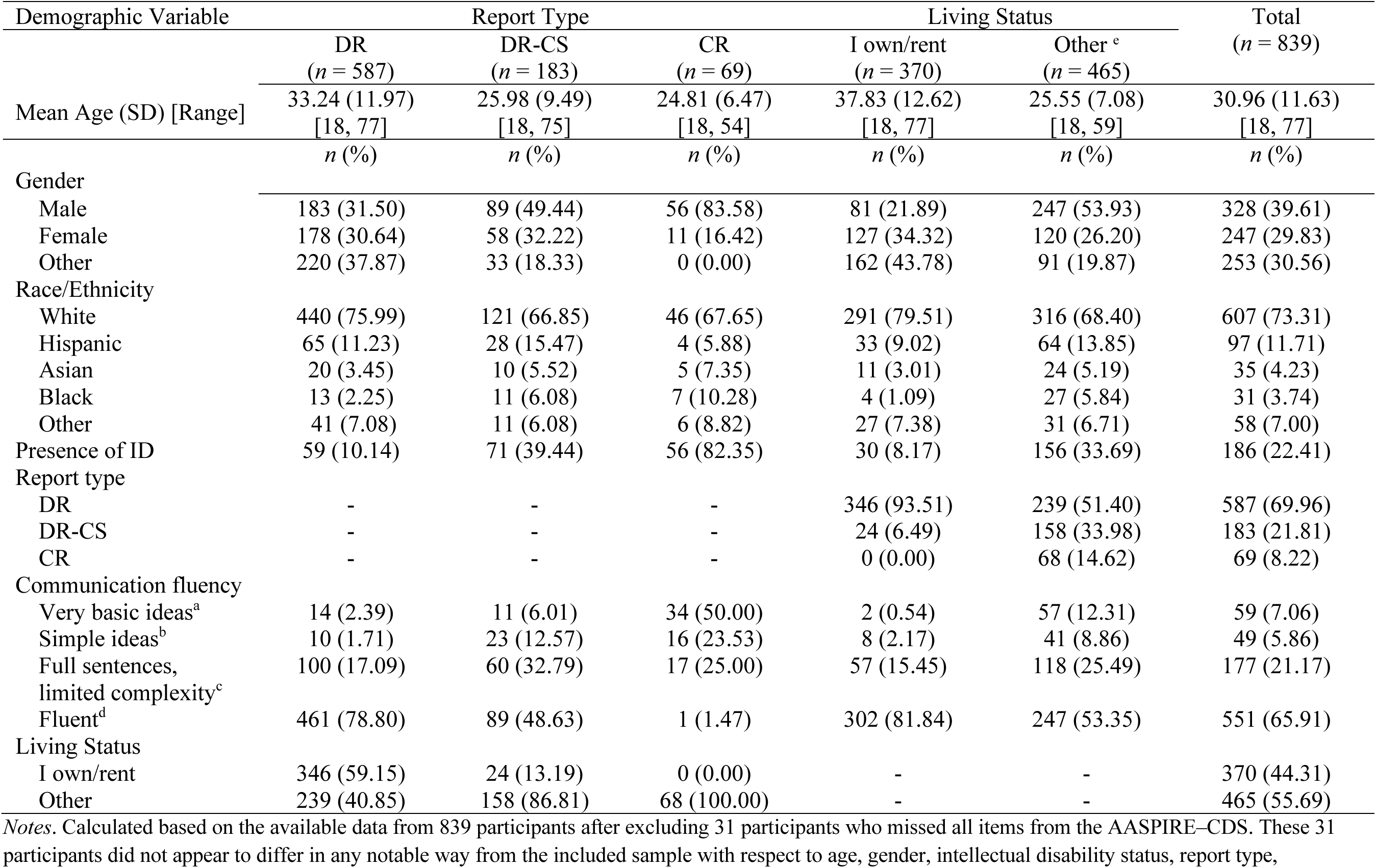
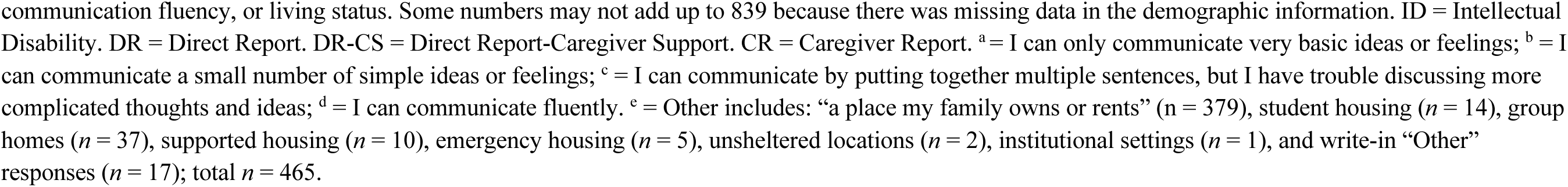
Demographic Characteristics.

| Demographic Variable | Report Type |  |  | Living Status |  | Total<br>(*n* = 839) |
| --- | --- | --- | --- | --- | --- | --- |
|  | DR<br>(*n* = 587) | DR-CS<br>(*n* = 183) | CR<br>(*n* = 69) | I own/rent<br>(*n* = 370) | Other <sup>e</sup><br>(*n* = 465) |  |
| Mean Age (SD) [Range] | 33.24 (11.97)<br>[18, 77] | 25.98 (9.49)<br>[18, 75] | 24.81 (6.47)<br>[18, 54] | 37.83 (12.62)<br>[18, 77] | 25.55 (7.08)<br>[18, 59] | 30.96 (11.63)<br>[18, 77] |
|  | *n* (%) | *n* (%) | *n* (%) | *n* (%) | *n* (%) | *n* (%) |
| Gender |  |  |  |  |  |  |
| Male | 183 (31.50) | 89 (49.44) | 56 (83.58) | 81 (21.89) | 247 (53.93) | 328 (39.61) |
| Female | 178 (30.64) | 58 (32.22) | 11 (16.42) | 127 (34.32) | 120 (26.20) | 247 (29.83) |
| Other | 220 (37.87) | 33 (18.33) | 0 (0.00) | 162 (43.78) | 91 (19.87) | 253 (30.56) |
| Race/Ethnicity |  |  |  |  |  |  |
| White | 440 (75.99) | 121 (66.85) | 46 (67.65) | 291 (79.51) | 316 (68.40) | 607 (73.31) |
| Hispanic | 65 (11.23) | 28 (15.47) | 4 (5.88) | 33 (9.02) | 64 (13.85) | 97 (11.71) |
| Asian | 20 (3.45) | 10 (5.52) | 5 (7.35) | 11 (3.01) | 24 (5.19) | 35 (4.23) |
| Black | 13 (2.25) | 11 (6.08) | 7 (10.28) | 4 (1.09) | 27 (5.84) | 31 (3.74) |
| Other | 41 (7.08) | 11 (6.08) | 6 (8.82) | 27 (7.38) | 31 (6.71) | 58 (7.00) |
| Presence of ID | 59 (10.14) | 71 (39.44) | 56 (82.35) | 30 (8.17) | 156 (33.69) | 186 (22.41) |
| Report type |  |  |  |  |  |  |
| DR | - | - | - | 346 (93.51) | 239 (51.40) | 587 (69.96) |
| DR-CS | - | - | - | 24 (6.49) | 158 (33.98) | 183 (21.81) |
| CR | - | - | - | 0 (0.00) | 68 (14.62) | 69 (8.22) |
| Communication fluency |  |  |  |  |  |  |
| Very basic ideas<sup>a</sup> | 14 (2.39) | 11 (6.01) | 34 (50.00) | 2 (0.54) | 57 (12.31) | 59 (7.06) |
| Simple ideas<sup>b</sup> | 10 (1.71) | 23 (12.57) | 16 (23.53) | 8 (2.17) | 41 (8.86) | 49 (5.86) |
| Full sentences,<br>limited complexity<sup>c</sup> | 100 (17.09) | 60 (32.79) | 17 (25.00) | 57 (15.45) | 118 (25.49) | 177 (21.17) |
| Fluent<sup>d</sup> | 461 (78.80) | 89 (48.63) | 1 (1.47) | 302 (81.84) | 247 (53.35) | 551 (65.91) |
| Living Status |  |  |  |  |  |  |
| I own/rent | 346 (59.15) | 24 (13.19) | 0 (0.00) | - | - | 370 (44.31) |
| Other | 239 (40.85) | 158 (86.81) | 68 (100.00) | - | - | 465 (55.69) |

Notes. Calculated based on the available data from 839 participants after excluding 31 participants who missed all items from the AASPIRE-CDS. These 31 participants did not appear to differ in any notable way from the included sample with respect to age, gender, intellectual disability status, report type,
communication fluency, or living status. Some numbers may not add up to 839 because there was missing data in the demographic information. ID = Intellectual Disability. DR = Direct Report. DR-CS = Direct Report-Caregiver Support. CR = Caregiver Report. <sup>a</sup> = I can only communicate very basic ideas or feelings; <sup>b</sup> = I can communicate a small number of simple ideas or feelings; <sup>c</sup> = I can communicate by putting together multiple sentences, but I have trouble discussing more complicated thoughts and ideas; <sup>d</sup> = I can communicate fluently. <sup>e</sup> = Other includes: “a place my family owns or rents” (*n* = 379), student housing (*n* = 14), group homes (*n* = 37), supported housing (*n* = 10), emergency housing (*n* = 5), unsheltered locations (*n* = 2), institutional settings (*n* = 1), and write-in “Other” responses (*n* = 17); total *n* = 465.

Participants completed surveys at three time points over a 12-18 month period, with the 2nd and 3rd time points 5-11 months and 12-18 months after the baseline, respectively. A 15% random sample subset of participants (*n* = 129) completed the survey again 2 weeks after Time 2 to assess test-retest reliability (details available in Nicolaidis et al., 2026b).

### Instruments

All scales are freely available at www.aaspire.org/measurement. Initial psychometric findings for the broader AASPIRE Measurement Toolkit are reported in Nicolaidis et al. (2026b).

#### AASPIRE-CDS and SDI:AR-AASPIRE

The 13-item AASPIRE-CDS asks whether individuals get to make decisions across a spectrum ranging from small, daily choices to major life decisions. For example, “I usually get to make my own decisions about my romantic or sexual relationships.” Response options use a 5-point Likert scale (1-Strongly disagree, 5-Strongly agree). A preface with clickable links for more information clarifies the focus on both opportunity and ability, opportunities for supported decision-making, and acknowledgement that greater opportunity to make choices may not always be desirable (see Supplementary Materials C for the full text provided in the clickable links).

The 21-item SDI:AR-AASPIRE represents individuals’ perceptions of autonomy (e.g., thinking about future possibilities), self-regulated action (e.g., setting own goals), and psychological empowerment (e.g., recognizing own strengths), rated on a 5-point Likert scale (1-Strongly disagree, 5-Strongly agree). We made minor adaptations to the SDI:AR by changing wording in a few items and adding a preface and hotlinks to clarify confusing terms, increase precision, and provide additional context that autistic partners felt was necessary. Internal consistency was high both excluding CR responses (*α* = 0.92, *ω* = 0.92) and including them (*α* = 0.93, *ω* = 0.93; see Supplementary Materials A for all changes and psychometric properties of the SDI:AR-AASPIRE). We also created a corresponding CR version of the SDI:AR-AASPIRE.

The DR versions of both AASPIRE-CDS and SDI:AR-AASPIRE included follow-up items about whether they received support and the type of support received to complete the survey. The CR versions included two closed-ended follow-up questions: how the caregiver knew the information and how confident they were in their responses (see Supplementary Tables S7 and S8, respectively, in Supplementary Materials D). Both DR and CR versions included a single item assessing perceived change over the past 6 months in the participant’s choices and decisions, asking whether “the things this survey asked about” had gotten worse, stayed about the same, or gotten better.

#### Grouping Variables for the Measurement Invariance Analyses

For the measurement invariance analyses, we classified participants into three report type categories: DR, DR-CS, and CR based on the survey type they selected during screening (DR or CR) and a question after each instrument asking if they received help while answering it (See Supplementary Table S9 in Supplementary Materials D for the types of support that DR-CS participants reported receiving). We measured living status using self- or proxy-reported answers to, “Which of the following best describes where you live?” Options included owning or renting one’s own place, living with family, student housing, group homes, institutional settings, and other arrangements (see Table 1). We recoded the living status variable into a binary format: Own Arrangement (“a place I own or rent”) versus Other due to convergence issues. The Pearson correlation between report type and living status was *r* = 0.39.

#### Measures Used to Assess Construct Validity

To assess AASPIRE–CDS’s convergent validity, we used items measuring living status (described above), communication fluency, personal care needs, independent living needs, and self-determination. We measured communication fluency with an item, “*Which of the following statements best describe your communication, under good conditions, using whatever works best for you (e.g., talking, writing, typing, sign language, gestures)?*” rated on a 4-point Likert scale (1-Very basic ideas or feelings, 2-Simple ideas or feelings, 3-Full Sentences with limited complexity, and 4-Fluent). We assessed personal care support needs with the item,*“How often do you need support to do any of your basic self-care activities such as eating, dressing, bathing, going to the bathroom, or brushing your teeth?”* We measured independent living needs with a parallel item focused on tasks such as cooking, cleaning, shopping, and managing money. Both were rated on a 3-point Likert scale (1-Rarely or never, 2-Sometimes, 3-Always or often).

We measured employment satisfaction using the AASPIRE Overall Employment Satisfaction Scale, which consists of ten items focused on aspects such as income, job fit, and career progress rated on a 5-point Likert scale (1-Strongly Disagree, 5-Strongly Agree). The scale has excellent internal consistency reliability (alpha = 0.91) and strong test-retest reliability (ICC 0.86) (Nicolaidis et al., 2026b). Additionally, we measured overall health with an item, “*In general, how would you rate your overall health*?” and assessed QoL with a general-purpose single-item rating of global QoL, “In general, how would you rate your QoL?” Both were rated on a 5-point Likert scale (1-Poor, 5-Excellent).

### Data Analysis

#### Missing data

Of the 839 participants included in the analysis, the proportion of missing responses per AASPIRE-CDS item ranged from 0.1% to 0.6% (affecting 1-5 participants per item), and 17 participants had at least one missing AASPIRE-CDS item. Among these 17 participants, 5 were CR respondents (29.4%) and 7 had intellectual disability (41.2%); in the full analytic sample, 69 participants were CR respondents (8.2%) and 186 had intellectual disability (22.4%). We therefore used multiple imputation to avoid disproportionately excluding participants with co-occurring intellectual disability and participants who needed caregivers to report on their behalf. We generated 10 imputed datasets using a conditional multivariate normal distribution based on observed response patterns. We conducted sensitivity analyses by repeating all analyses using pairwise deletion instead of imputation, which showed patterns of results consistent with the main results we report in this manuscript.

#### Identifying the Factor Structure and Measurement Invariance

To identify AASPIRE–CDS’s underlying structure, we randomly split the dataset in half, stratified by report type. We conducted an EFA on one half of the data using principal axis factoring with varimax rotation and Kaiser normalization. We used the same split-half sample from the EFA to conduct Horn’s parallel analysis with 5,000 iterations and the 95th percentile criterion. We retained factors when the observed eigenvalue exceeded the corresponding random-data eigenvalue. We used the EFA and parallel analysis results to identify the initial factor structure.

We then used confirmatory factor analysis (CFA) to test this structure on the second half of the sample. We evaluated CFA model fit using the following criteria: root-mean-square error of approximation (RMSEA) ≤ 0.09, comparative fit index (CFI) and Tucker-Lewis index (TLI) ≥ 0.90, and standardized root-mean-square residual (SRMR) < 0.08 (Brown, 2015; Little, 2013). We performed a sensitivity check by iteratively freeing the largest modification index at each step and repeating the process until identifying a well-fitting model (Byrne et al., 1989). Finally, we evaluated the standardized factor loadings, considering loadings ≥ 0.30 salient (Brown, 2015).

To test measurement invariance, we used the full dataset to ensure model convergence. We first evaluated the model fits separately within each subgroup of the grouping variables and conducted multigroup CFAs to test measurement invariance for report type and living status. For measurement invariance analyses involving living status, we conducted the full set of analyses both excluding and including the CR group. We then evaluated measurement invariance in three sequential steps for each variable: configural (baseline), metric (weak), and scalar (strong). Configural invariance tests whether the factor structure is consistent across groups. Metric invariance assesses whether different groups interpret the construct similarly. Scalar invariance determines whether groups use the response scale comparably. We evaluated the addition of invariance constraints by comparing nested models and considered the more constrained model tenable when the decrease in CFI was less than or equal to 0.01, indicating that added constraints did not substantially worsen model fit (Cheung & Rensvold, 2002). We also calculated Akaike’s Information Criterion (AIC) as a supplementary model-comparison index for the configural, metric, scalar, and partial scalar models. We interpreted AIC descriptively, with lower values indicating better relative fit among models estimated with the same analytic sample. When full metric or scalar invariance was not supported, we pursued partial invariance by inspecting modification indices and relaxing equality constraints one at a time for parameters with significant misfit (p < 0.05). After establishing sufficient invariance, we used Wald tests as post-hoc analyses to examine group differences in latent means and variances.

In parallel, we applied the alignment optimization method to assess approximate invariance, following recommendations by Asparouhov and Muthén (2014) and Marsh et al. (2018). We conducted all analyses in Stata, except for alignment optimization, which we implemented in R using the lavaan package.

We also conducted secondary measurement invariance analyses by autistic participants’ gender, subcohort, and communication fluency. Full or partial measurement invariance was achieved for all grouping variables. See Supplementary Materials E for the rationale for these secondary analyses, demographic characteristics by grouping variables (Supplementary Table S10), CFA and measurement invariance results (Supplementary Table S11), and Wald tests of latent mean and variance differences (Supplementary Table S12).

#### Internal Consistency

To examine internal consistency, we calculated coefficient alpha and coefficient omega using datasets that included and excluded CR. Additionally, we calculated internal consistency separately within the CR group. We assessed test-retest reliability by calculating intraclass correlation coefficients (McGraw & Wong, 1996; Shrout & Fleiss, 1979) between Time 2 scores and those collected 2 weeks later.

#### Responsiveness to Change

We calculated two interval-specific change scores by subtracting mean AASPIRE-CDS scores at Time 1 from Time 2 and Time 2 from Time 3, with positive scores indicating improvement and negative scores indicating worsening in opportunities to make choices and decisions. We used linear regression with robust standard errors clustered by participant to examine whether change in mean AASPIRE-CDS scores differed across a single self-reported change variable with three categories (improved, stayed the same, worsened), with “stayed the same” specified as the reference category.

#### Construct Validity

We conducted Pearson correlations including AASPIRE–CDS, SDI:AR-AASPIRE, and all validity indicators for both instruments. Supplementary Materials F reports assumption checks for the Pearson correlation analyses. Additionally, we conducted tests of the equality of correlation coefficients (using the cortesti command) to examine whether the correlations between each construct validity indicator and SDI:AR-AASPIRE differed significantly from the corresponding correlations between the same construct validity indicator and AASPIRE–CDS. We repeated all analyses after excluding the CR group and also conducted the same construct validity analyses within the CR group. All analyses used α = 0.05; we did not apply multiple-comparison corrections.

## Results

### Identifying the Factor Structure and Measurement Invariance

In the initial EFA conducted on half of the dataset, we extracted two factors based on the eigenvalue > 1 criterion. The model explained 58.66% of the total variance, with Factors 1 and 2 yielding eigenvalues of 6.58 and 1.05, respectively. The Kaiser–Meyer–Olkin was 0.94. Several items exhibited cross-loadings with similarly strong loadings on both factors. Given the conceptual overlap among items and the fact that the scale was designed to capture a single gradient of decision-making rather than distinct subdimensions, we re-examined the eigenvalues and scree plot. The large drop between the first and second eigenvalues and the clear elbow in the scree plot both indicated a strongly dominant first factor, supporting a unidimensional structure. We therefore retained a single-factor structure and re-specified the EFA using one factor. All 13 items loaded strongly onto this factor, with loadings ranging from 0.59 to 0.78. The model explained 50.58% of the total variance, and Factor 1 had an eigenvalue of 6.58, supporting the unidimensionality of AASPIRE–CDS.

Horn’s parallel analysis also supported retaining one factor. The first observed eigenvalue exceeded the corresponding 95th percentile random-data eigenvalue, 6.58 versus 1.37, yielding a positive adjusted eigenvalue of 5.21. Although the second observed eigenvalue was slightly greater than 1.00, it did not exceed the corresponding random-data eigenvalue, 1.05 versus 1.28, yielding a negative adjusted eigenvalue of -0.22. All subsequent adjusted eigenvalues were also negative.

We conducted a CFA on the second half of the data to confirm the one-factor structure. The initial unidimensional model yielded RMSEA = 0.109, CFI = 0.881, TLI = 0.857, and SRMR = 0.055. After we iteratively added 14 error covariances, the model yielded RMSEA = 0.078, CFI = 0.952, TLI = 0.926, and SRMR = 0.039, which met the predefined fit criteria. Standardized factor loadings ranged from 0.55 to 0.78, and differed only minimally from the loadings in the initial model without correlated errors.

In separate CFAs conducted within each level of the grouping variables in the combined sample, the DR and DR-CS models met the predefined fit criteria, whereas the CR model did not meet all criteria (Table 2). Therefore, we conducted the subsequent measurement invariance for report type after excluding the CR group. To ensure this exclusion did not alter the core structure, we re-ran the EFA and CFA without the CR group, and the overall pattern of results remained consistent. Supplementary Materials G reports sensitivity analyses examining the AASPIRE-CDS factor structure with and without the CR group, including the initial EFA results (Supplementary Table S13), the re-specified one-factor EFA results (Supplementary Table S14), CFA standardized loadings (Supplementary Table S15), and the scree plot excluding the CR group (Supplementary Figure S1).

**Table 2.**
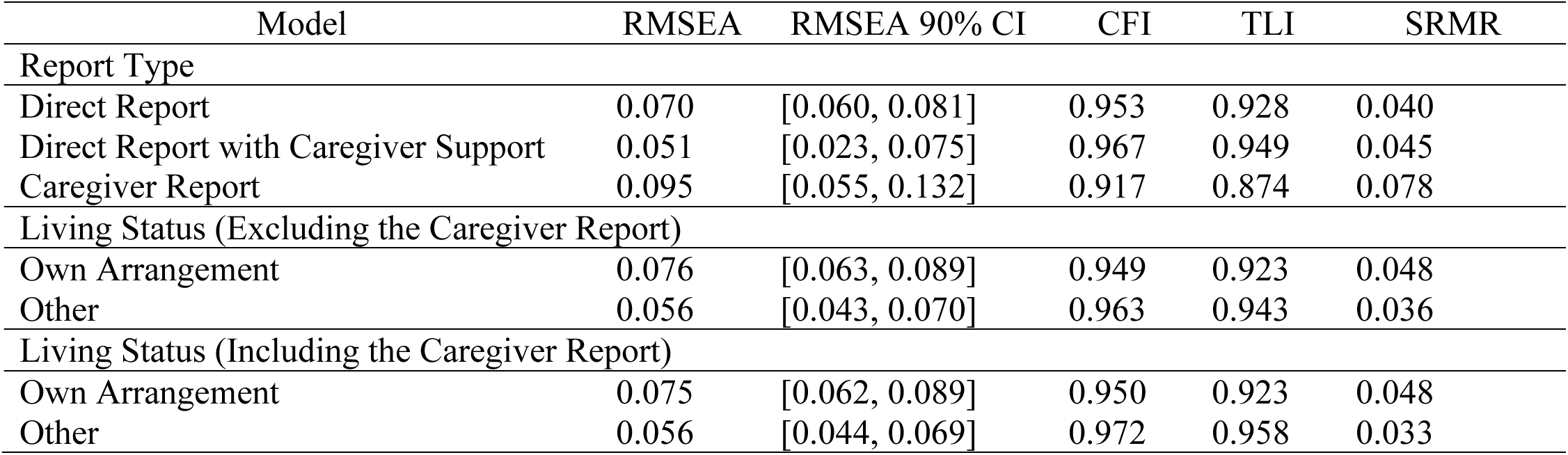
CFA Fit Indices Within Report Type and Living Status.

| Model | RMSEA | RMSEA 90% CI | CFI | TLI | SRMR |
| --- | --- | --- | --- | --- | --- |
| Report Type |  |  |  |  |  |
| Direct Report | 0.070 | [0.060, 0.081] | 0.953 | 0.928 | 0.040 |
| Direct Report with Caregiver Support | 0.051 | [0.023, 0.075] | 0.967 | 0.949 | 0.045 |
| Caregiver Report | 0.095 | [0.055, 0.132] | 0.917 | 0.874 | 0.078 |
| Living Status (Excluding the Caregiver Report) |  |  |  |  |  |
| Own Arrangement | 0.076 | [0.063, 0.089] | 0.949 | 0.923 | 0.048 |
| Other | 0.056 | [0.043, 0.070] | 0.963 | 0.943 | 0.036 |
| Living Status (Including the Caregiver Report) |  |  |  |  |  |
| Own Arrangement | 0.075 | [0.062, 0.089] | 0.950 | 0.923 | 0.048 |
| Other | 0.056 | [0.044, 0.069] | 0.972 | 0.958 | 0.033 |

The measurement invariance model for report type in the combined DR and DR-CS sample, after removing the CR group, met the fit criteria, with full configural and metric invariance supported and partial scalar invariance achieved by freeing the intercepts of two items (Health and Outside Home; Table 3). The model fit within each subgroup of living status met the fit criteria both when including and excluding the CR group (Table 2). We also established configural and metric invariance and achieved partial scalar invariance by freeing the intercepts of *Health*, *Communicate*, and *Outside Home* both excluding and including the CR (Table 3). Across both report-type and living-status invariance analyses, the AIC values reported in Table 3 suggested an interpretation consistent with that based on the ΔCFI-based results.

**Table 3.** Fit Indices for Multiple-Group CFA.

| Model | RMSEA | RMSEA 90% CI | CFI | TLI | SRMR | Tenable | ΔCFI | AIC | Variables freed |
| --- | --- | --- | --- | --- | --- | --- | --- | --- | --- |
| Report Type (Direct Report vs. Direct Report with Caregiver Support) Excluding the Caregiver Report |  |  |  |  |  |  |  |  |  |
| Configural | 0.065 | [0.055, 0.074] | 0.957 | 0.935 | 0.042 | N/A | N/A | 22842.46 | - |
| Metric | 0.060 | [0.051, 0.069] | 0.959 | 0.944 | 0.050 | Yes | 0.002 | 22824.96 | - |
| Scalar | 0.067 | [0.058, 0.075] | 0.944 | 0.931 | 0.053 | No | -0.015 | 22869.77 | - |
| Scalar (P) | 0.062 | [0.053, 0.071] | 0.953 | 0.941 | 0.052 | Yes | -0.006 | 22837.48 | Health, Outside Home |
| Living Status (Own Arrangement vs. Other) Excluding the Caregiver Report |  |  |  |  |  |  |  |  |  |
| Configural | 0.066 | [0.057, 0.076] | 0.955 | 0.932 | 0.042 | N/A | N/A | 22585.13 | - |
| Metric | 0.064 | [0.055, 0.073] | 0.954 | 0.937 | 0.053 | Yes | -0.001 | 22579.95 | - |
| Scalar | 0.075 | [0.066, 0.083] | 0.930 | 0.914 | 0.057 | No | -0.024 | 22657.70 | - |
| Scalar (P) | 0.067 | [0.059, 0.076] | 0.944 | 0.929 | 0.055 | Yes | -0.010 | 22611.36 | Communicate, Outside Home, Health |
| Living Status (Own Arrangement vs. Other) Including the Caregiver Report |  |  |  |  |  |  |  |  |  |
| Configural | 0.065 | [0.056, 0.074] | 0.962 | 0.943 | 0.041 | N/A | N/A | 25380.13 | - |
| Metric | 0.064 | [0.055, 0.071] | 0.960 | 0.945 | 0.054 | Yes | -0.002 | 25380.04 | - |
| Scalar | 0.074 | [0.066, 0.082] | 0.940 | 0.926 | 0.062 | No | -0.020 | 25465.05 | - |
| Scalar (P) | 0.068 | [0.059, 0.076] | 0.952 | 0.939 | 0.058 | Yes | -0.008 | 25411.60 | Communicate, Outside Home, Health |

*Notes.* (P) = Partial Invariance Model. N/A = Not applicable. AIC = Akaike's Information Criterion. ΔCFI represents the change in CFI from the comparison model to the focal model. Positive values indicate an increase in CFI, and negative values indicate a decrease in CFI. Partial scalar models were compared with the metric model. We considered models tenable when CFI decreased by no more than 0.01.

The alignment results further supported the tentative tenability of measurement invariance, as no intercepts were flagged as non-invariant in any model across the grouping variables, whether the CR group was excluded or included. Overall, we tentatively accepted measurement invariance for AASPIRE–CDS across report type after excluding CR and across living status in analyses both including and excluding the CR group.

Table 4 presents the Wald test results comparing latent means and variances across the grouping variables: for report type (after excluding the CR group) and for living status (including and excluding CR).

**Table 4.** Wald Test Results for Group Differences in Latent Means and Variances.

| Latent Variable | Groups | Latent Mean | $\chi^2$ | $p$-value | Latent Variances | $\chi^2$ | $p$-value |
| --- | --- | --- | --- | --- | --- | --- | --- |
| Report Type | Direct Report | 4.24 | 33.06 | < 0.0001 | 0.40 | 0.38 | 0.54 |
|  | Direct Report with Caregiver Support | 3.88 |  |  | 0.44 |  |  |
| Living Status<br>(Excluding the CR) | Own Arrangement | 4.32 | 43.94 | < 0.0001 | 0.35 | 3.80 | 0.05 |
|  | Other | 3.99 |  |  | 0.44 |  |  |
| Living Status<br>(Including the CR) | Own Arrangement | 4.32 | 90.57 | < 0.0001 | 0.36 | 28.94 | < 0.0001 |
|  | Other | 3.78 |  |  | 0.70 |  |  |

### Internal Consistency

AASPIRE–CDS, excluding CR, demonstrated high internal consistency reliability, with both Cronbach’s alpha and McDonald’s omega equal to 0.90; when including CR, both Cronbach’s alpha and McDonald’s omega were 0.92. Within the CR group, AASPIRE–CDS demonstrated internal consistency with α = 0.88 and ω = 0.89. Test-retest reliability was 0.95.

### Responsiveness to Change

The self-reported change in the construct assessed category was significantly associated with change in mean AASPIRE-CDS scores, F(2, 753) = 18.74, *p* < 0.001, *R*² = .029. Compared with participants who reported that their opportunities to make choices stayed the same, those who reported improved opportunities to make choices had more positive change in mean CDS scores (*b* = 0.12, *p* = 0.001), whereas those who reported worsening opportunities had significantly more negative change in mean CDS scores (*b* = -0.31, *p* < 0.001).

## Construct Validity

Table 5 depicts the results of Pearson correlation analyses for the full sample involving AASPIRE–CDS, SDI:AR-AASPIRE, and their associated construct validity indicators, results of the equality of correlation coefficients comparing these correlations, and the corresponding correlation results calculated including and excluding CR. Correlations within the CR group were generally weaker than those observed in the combined DR and DR-CS groups. Figures 1 and 2 present associations of AASPIRE-CDS and SDI:AR-AASPIRE with AASPIRE-CDS convergent validity variables and SDI:AR-AASPIRE convergent validity variables, respectively.

**Table 5.**
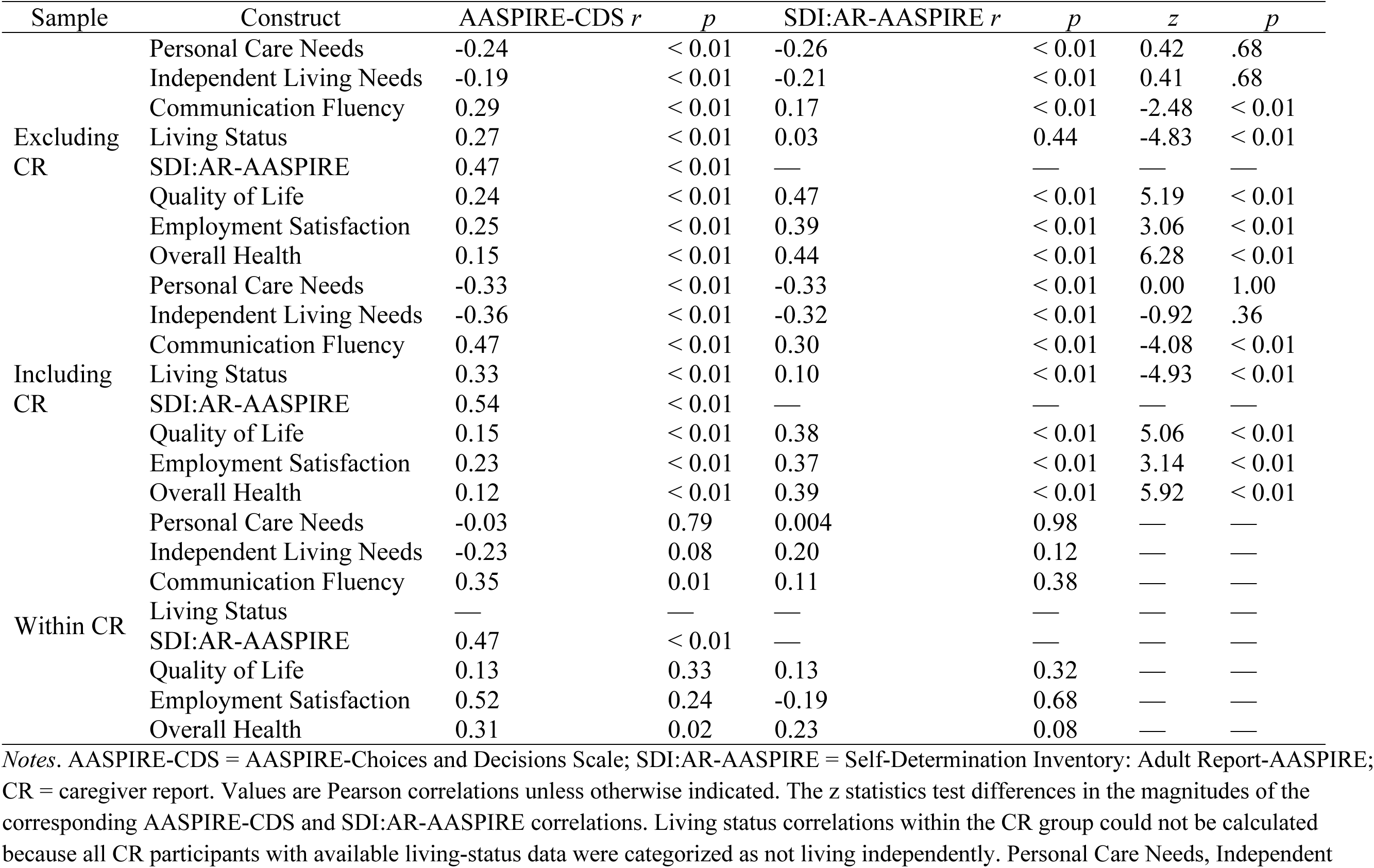
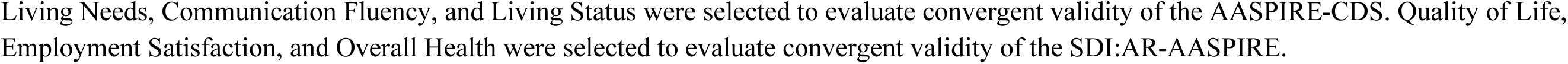
Construct Validity Correlations.

| Sample | Construct | AASPIRE-CDS *r* | *p* | SDI:AR-AASPIRE *r* | *p* | *z* | *p* |
| --- | --- | --- | --- | --- | --- | --- | --- |
| Excluding CR | Personal Care Needs | -0.24 | < 0.01 | -0.26 | < 0.01 | 0.42 | .68 |
|  | Independent Living Needs | -0.19 | < 0.01 | -0.21 | < 0.01 | 0.41 | .68 |
|  | Communication Fluency | 0.29 | < 0.01 | 0.17 | < 0.01 | -2.48 | < 0.01 |
|  | Living Status | 0.27 | < 0.01 | 0.03 | 0.44 | -4.83 | < 0.01 |
|  | SDI:AR-AASPIRE | 0.47 | < 0.01 | — | — | — | — |
|  | Quality of Life | 0.24 | < 0.01 | 0.47 | < 0.01 | 5.19 | < 0.01 |
|  | Employment Satisfaction | 0.25 | < 0.01 | 0.39 | < 0.01 | 3.06 | < 0.01 |
|  | Overall Health | 0.15 | < 0.01 | 0.44 | < 0.01 | 6.28 | < 0.01 |
| Including CR | Personal Care Needs | -0.33 | < 0.01 | -0.33 | < 0.01 | 0.00 | 1.00 |
|  | Independent Living Needs | -0.36 | < 0.01 | -0.32 | < 0.01 | -0.92 | .36 |
|  | Communication Fluency | 0.47 | < 0.01 | 0.30 | < 0.01 | -4.08 | < 0.01 |
|  | Living Status | 0.33 | < 0.01 | 0.10 | < 0.01 | -4.93 | < 0.01 |
|  | SDI:AR-AASPIRE | 0.54 | < 0.01 | — | — | — | — |
|  | Quality of Life | 0.15 | < 0.01 | 0.38 | < 0.01 | 5.06 | < 0.01 |
|  | Employment Satisfaction | 0.23 | < 0.01 | 0.37 | < 0.01 | 3.14 | < 0.01 |
|  | Overall Health | 0.12 | < 0.01 | 0.39 | < 0.01 | 5.92 | < 0.01 |
| Within CR | Personal Care Needs | -0.03 | 0.79 | 0.004 | 0.98 | — | — |
|  | Independent Living Needs | -0.23 | 0.08 | 0.20 | 0.12 | — | — |
|  | Communication Fluency | 0.35 | 0.01 | 0.11 | 0.38 | — | — |
|  | Living Status | — | — | — | — | — | — |
|  | SDI:AR-AASPIRE | 0.47 | < 0.01 | — | — | — | — |
|  | Quality of Life | 0.13 | 0.33 | 0.13 | 0.32 | — | — |
|  | Employment Satisfaction | 0.52 | 0.24 | -0.19 | 0.68 | — | — |
|  | Overall Health | 0.31 | 0.02 | 0.23 | 0.08 | — | — |

*Notes.* AASPIRE-CDS = AASPIRE-Choices and Decisions Scale; SDI:AR-AASPIRE = Self-Determination Inventory: Adult Report-AASPIRE; CR = caregiver report. Values are Pearson correlations unless otherwise indicated. The *z* statistics test differences in the magnitudes of the corresponding AASPIRE-CDS and SDI:AR-AASPIRE correlations. Living status correlations within the CR group could not be calculated because all CR participants with available living-status data were categorized as not living independently. Personal Care Needs, Independent
Living Needs, Communication Fluency, and Living Status were selected to evaluate convergent validity of the AASPIRE-CDS. Quality of Life, Employment Satisfaction, and Overall Health were selected to evaluate convergent validity of the SDI:AR-AASPIRE.

**Figure 1.**
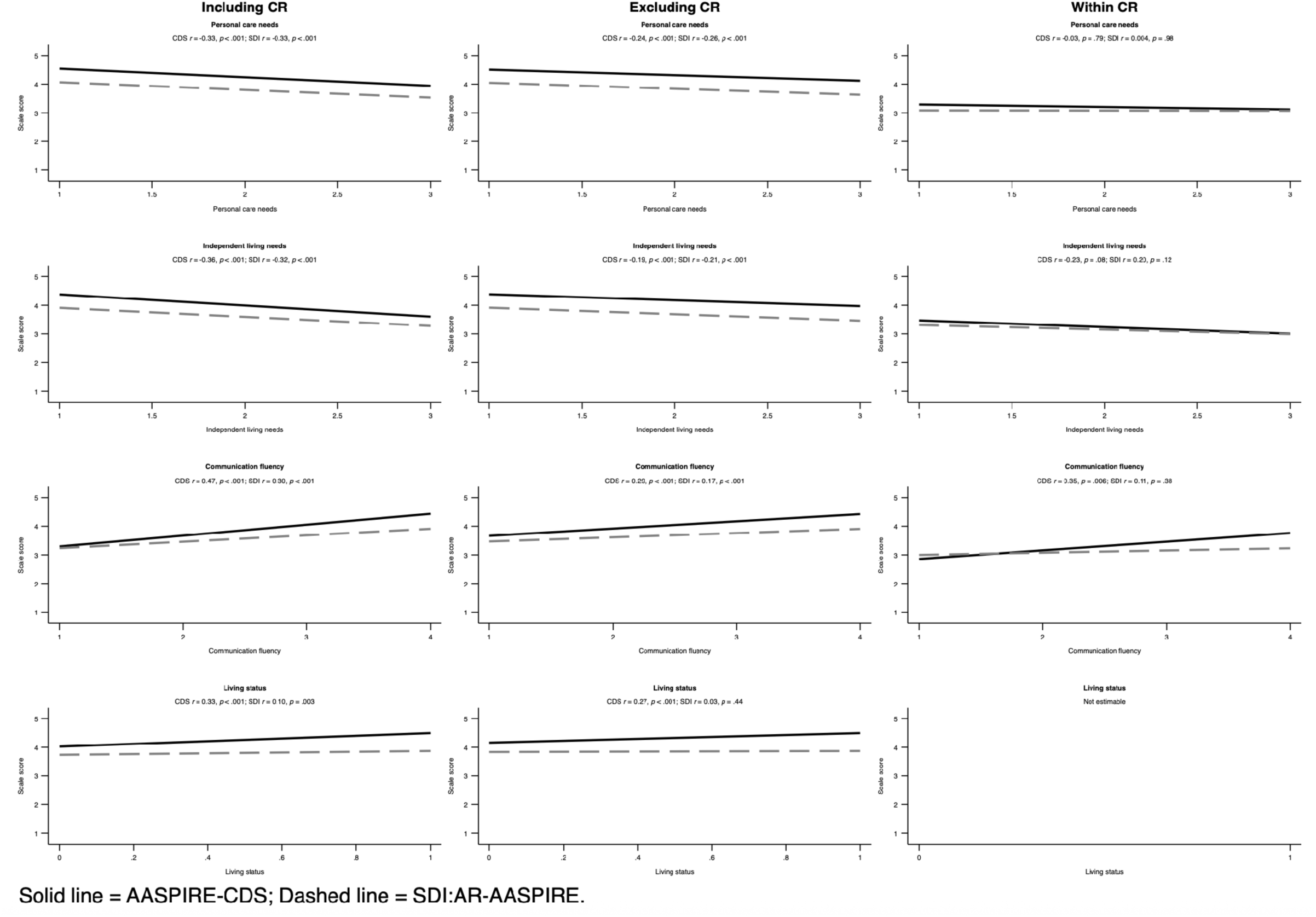
Associations of AASPIRE-CDS and SDI:AR-AASPIRE with Convergent Validity Variables for AASPIRE-CDS

**Figure 2.**
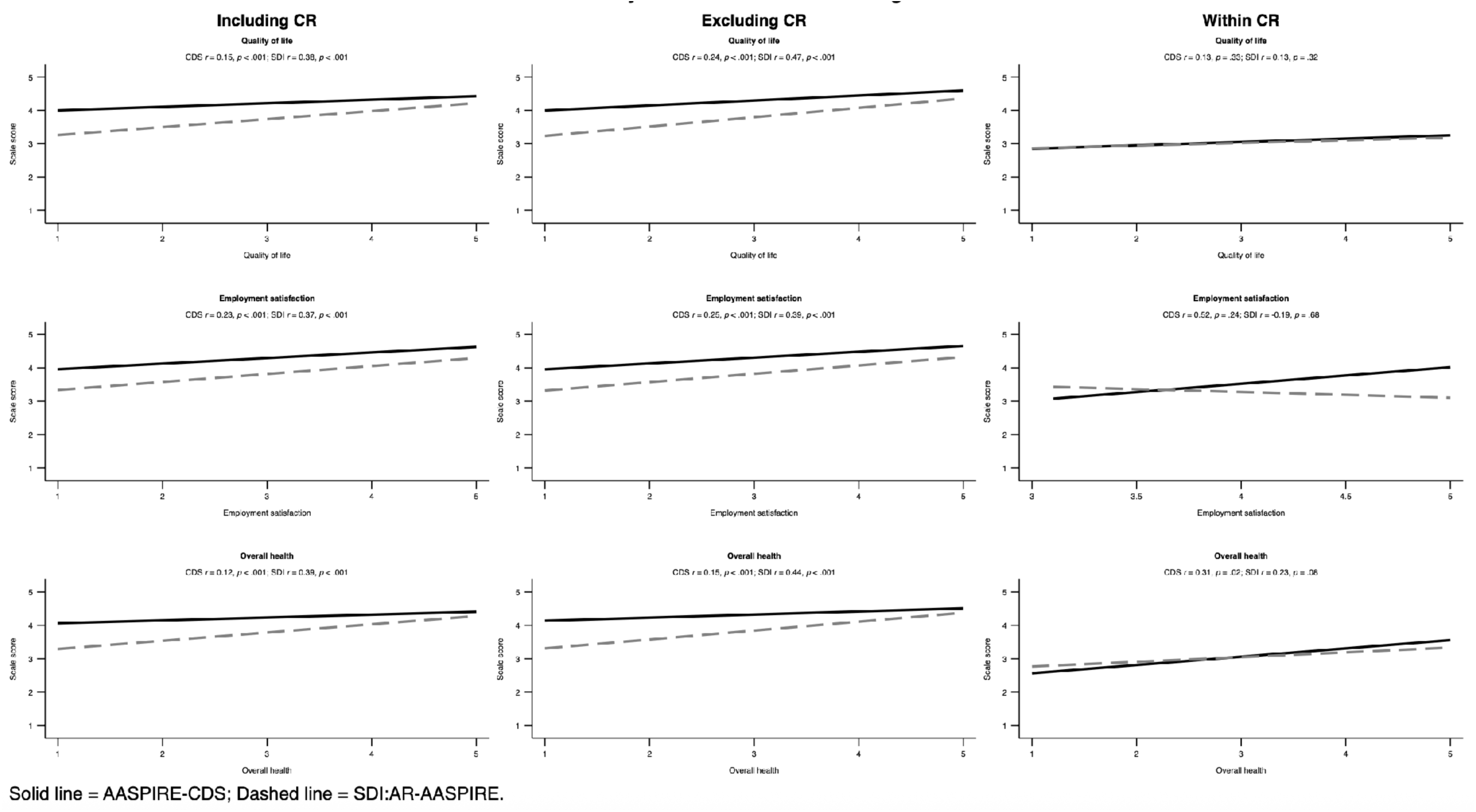
Associations of AASPIRE-CDS and SDI:AR-AASPIRE with Convergent Validity Variables for SDI:AR-AASPIRE

## Discussion

This study provides psychometric evidence that AASPIRE–CDS is a valid instrument with high internal consistency and test-retest reliabilities for measuring opportunities for making choices and decisions in autistic adults across different support needs and contexts.

### Factor Structure and Measurement Invariance of AASPIRE–CDS

In contrast to other instruments like NCI-IDD (HSRI & NASDDDS, 2025), which often measure individual choices as an indicator of formal developmental services quality and separate choices into multiple subdomains, AASPIRE–CDS assesses choice as a self-reported outcome and appears to reflect a unidimensional construct. This may be because we intentionally created items along a continuum representing increasingly complex or consequential choices and decisions.

AASPIRE–CDS demonstrated measurement invariance across living status groups, both with and without CR data; however, invariance for report type was achieved only after excluding CR data. This may be because caregivers interpret and respond to items differently from autistic individuals, consistent with prior research showing that proxy reports can diverge from self-reports when assessing subjective or non-observable experiences (Loftus et al., 2022; Ward et al., 2025). Other work has also suggested that self- and proxy-report discrepancies may reflect differential interpretation of items (Stokes et al., 2017). Minor wording differences in the CR version and/or the follow-up questions about caregivers’ information sources and confidence in their ratings, which were only present in the CR survey, may also have contributed to these discrepancies. It is also notable that latent variances differed substantially across living status only when CR data were included, with larger variances appearing in the non-independent living group. Because all CR participants were in the non-independent living group, this pattern may partly reflect the inclusion of CR data only in the non-independent living category, especially given that the CR-specific CFA did not meet the predefined model-fit criteria.

Latent mean comparisons indicated that autistic adults who completed the survey independently and who lived independently reported greater opportunities to make choices and decisions than their counterparts. This finding for living status also aligns with prior research showing that congregate or institutional settings living offers less opportunity for autonomous decisions (Kishi et al., 1988; Kozma et al., 2009; Tichá et al., 2012; Wehmeyer & Metzler, 1995). However, we did not directly assess impacts of congregate or institutional settings, given limited representation of participants in these settings.

### Convergent and Discriminant Validity

AASPIRE–CDS and SDI:AR-AASPIRE were moderately positively correlated, consistent with the expectation that they assess related but distinct constructs. As hypothesized, AASPIRE–CDS was more strongly associated with some variables known to impact available choices (communication fluency and independent living status) than SDI:AR-AASPIRE was associated with those variables, both when CR data were included and excluded, further supporting construct validity of the AASPIRE-CDS. Unexpectedly, personal care and independent living needs were similarly correlated with both AASPIRE-CDS and SDI:AR-AASPIRE. This may be because needing more support with everyday activities can limit some choices when support is unavailable or controlled by others, and repeated experiences of restricted choice may in turn shape perceived agency to exercise self-determination. This interpretation is consistent with Taub and Werner (2023), who emphasize that support needs do not necessarily undermine self-determination; rather, the availability and quality of support may determine whether individuals can make and act on their own choices.

Consistent with hypotheses, correlations between AASPIRE–CDS and well-being indicators were consistently weaker than those for SDI:AR-AASPIRE, confirming discriminant validity from internal self-determination as measured by SDI:AR-AASPIRE. Nonetheless, AASPIRE–CDS demonstrated associations with these well-being indicators too. This finding is consistent with evidence that choices promote QoL for people with intellectual disability (Brown & Brown, 2009; Neely-Barnes, 2008), although it could also be an artifact of the correlation between SDI:AR-AASPIRE and AASPIRE-CDS.

In the CR only group, almost no associations between AASPIRE-CDS, SDI:AR-AASPIRE and validity indicators were observed; SDI:AR-AASPIRE was unrelated to any outcomes and AASPIRE-CDS was only related to communication fluency and overall health. Differences between the DR/DR-CS and CR groups echo the broader finding that CR data function differently psychometrically. The small CR sample and heterogeneity in types of support may also have limited our power to detect associations.

Beyond the evidence for factor structure and construct validity, AASPIRE–CDS demonstrated strong content validity through CBPR-driven development and cognitive interviews, high internal consistency, and test–retest reliability. Further, we found that AASPIRE-CDS scores changed in the expected direction relative to self-reported changes in the construct being assessed by the AASPIRE-CDS over time, providing preliminary evidence of sensitivity to perceived change.

### Limitations and Future Directions

There are several limitations to consider. First, the extent to which the AASPIRE-CDS is best conceptualized as a single dimension or as two partially overlapping dimensions was not fully resolved, and future replication studies should revisit its factor structure. Second, as in most self-determination research, this study relies on reported self-determination rather than direct observation of choice-making processes. Future observational work is needed to clarify the extent to which self-reported self-determination corresponds to actual choice-making behavior. Third, we did not assess measurement invariance across time, which is an essential future direction for longitudinal work given that choice measures may function differently at different points in time (Lineberry et al., 2025). Evidence of longitudinal measurement invariance is needed to meaningfully compare data across time points (Vandenberg & Lance, 2000). Relatedly, the responsiveness-to-change finding should be interpreted with caution because it explained a small amount of variance and relied on a single perceived-change item. Future longitudinal studies should test whether the AASPIRE-CDS detects changes using independently measured indicators or in response to interventions or service changes.

Fourth, although we collected caregiver response confidence in the CR group and information about the type of support participants received in the DR-CS group, the small number of CRs and the wide variability in support types limited our ability to examine how these factors influenced response patterns (see Supplementary Materials D for descriptive statistics on caregivers’ confidence in their CR responses and type of support). Further, differences in caregiver roles (e.g., parent [Ferguson et al., 2025] vs. paid staff [Ryan et al., 2025]) could have influenced patterns, but we were unable to examine these differences because of the small caregiver-report sample and limited representation across caregiver roles. This sample-size limitation also precluded stable differential item functioning (DIF) analyses, as approximately 200 participants per group are often recommended for scales with more than two items (Scott et al., 2009). We also did not collect information about caregivers’ autism status. There are instances in which self-report may not be feasible for some autistic adults, and researchers may still choose to use CR in some contexts; however, caution is warranted when using and interpreting CR as a substitute for autistic self-report.

Future studies should include larger CR samples with contextual information (e.g., type of supports received, caregiver role, caregiver autism status) to better understand when and how CR data can be meaningfully interpreted and to examine how item-level response patterns may differ by report type, including through Rasch-based DIF analyses. A more diverse sample would allow further examination of how specific living arrangements (e.g., supported housing, group homes), communication styles (e.g., AAC use, intermittent speech: Bottema-Beutel et al., 2025; Zisk, 2024), and race and ethnicity (Girolamo et al., 2024; Song et al., 2026) may relate to self-determination and decision-making opportunities. These analyses were not feasible in the current study due to limited representation of key groups.

Fifth, although we examined correlations between AASPIRE-CDS scores and support-related indicators as part of convergent validity testing, these analyses were bivariate, as is the norm when assessing for convergent validity (e.g., Campbell & Fiske, 1959; Duckworth & Kern, 2011). We did not consider these indicators in relation to AASPIRE-CDS simultaneously because doing so would assess different research questions than ours. Future studies should use multivariable approaches to examine which of these factors are uniquely associated with opportunities to make choices and decisions, both concurrently and longitudinally, as well as how these factors may operate together.

Finally, we did not examine the external conditions that enable choice-making, which remain understudied. There is a clear need for more work examining how relational (e.g., caregiver expectations) or contextual (e.g., access to communication devices and community) factors shape internal perceptions of agency, external opportunities to enact choices, and collective self-determination (e.g., neurodiversity advocacy) among autistic adults across diverse contexts. Since independence, autonomy, and interdependence are defined and valued differently across cultures (Moza et al., 2021; Woodfield et al., 2025), cross-cultural research is needed to explore how choice opportunity structures vary by cultural expectations, service systems, and family roles, and whether measures function similarly across settings. Similarly, expectations and needs related to independence and autonomy vary across the lifespan, emphasizing the importance of considering how autistic adults’ access to opportunities to make choices and decisions may change as they age, and the corresponding impact on their self-determination and overall well-being. Future research should also investigate how alignments or mismatches between individual self-determination and external opportunities influence long-term outcomes such as QoL.

## Implications

This study provides evidence that AASPIRE–CDS and the SDI:AR-AASPIRE reliably and validly capture distinct yet complementary dimensions of self-determination in autistic adults, particularly when collecting data from autistic individuals. While internal psychological dimensions of self-determination have been the focus of considerable research (e.g., Kim, 2019; Morán et al., 2021), the external conditions that enable choice-making remain understudied. In this study, autistic adults who completed the survey independently and those living independently reported greater opportunities to make choices and decisions than their counterparts. Autistic people, particularly those with higher support needs, may require accommodations such as additional processing time, as well as opportunities to make mistakes with support, in order to exercise choice and build self-determination. Accordingly, supported decision-making should be available across services, families, and institutions (McDonald et al., 2021), alongside structured training to build the competence and commitment needed to support decision-making for autistic adults who require assistance. Findings also highlight needed policy-level actions including requiring training to promote autistic agency in making choices across service systems, guaranteeing access to usable communication for decision-making (with AAC available when needed; Srinivasan et al., 2025), and strengthening the assessment of choice and decision opportunities in both service quality evaluation and research.

## Data Availability

All data from this study will be available through the National Institutes of Health (NIH) National Data Archive (NDA).

## Acknowledgements

We would like to thank other members of the AASPIRE Outcomes Project team, including Shannon Des Roches Rosa, Todd Edwards, Clarissa Kripke, Julia Love, and Julie Lounds Taylor, for their contributions to creating study materials, and K.J. Flores for their help with participant recruitment and data collection.

## Ethical considerations

The Institutional Review Board (IRB) at Portland State University (PSU) approved the project and served as the Single IRB (approval # 227674-18); Oregon Health & Science University, Vanderbilt University Medical Center, and San Diego State University waived authorization to the PSU IRB. Involvement by the Multnomah County Developmental Disability Services was subsumed under the PSU IRB.

## Consent to participate

Participants or their legally authorized representatives gave written consent online or in-person.

