## Supplementary Materials for "Self- and Caregiver-Reported Choice-Making in Autistic Adults: Development and Validation of the AASPIRE–Choices and Decisions Scale"

**Supplementary Materials A**

#### Adaptation and Psychometric Properties of the SDI:AR-AASPIRE

#### List of Changes Made to the SDI:AR-AASPIRE



### Detailed Comparison Between Original and Adapted Version

Changes between the original Self-Determination Inventory and the Self-Determination Inventory – AASPRIE

| | Original | Revised |
|---|---|---|
| **Introduction** | 1. Read each item.<br><br>2. Click or touch the place on the line or use the left and right arrow keys to show how much you agree or disagree. The darker the line (or the higher the number for those using a screen reader), the more you agree | Being self-determined is about making things happen in your life, with as many supports as you want or need.<br><br>The next set of questions focuses on how much:<br><br>• You feel that you end up making choices and decisions that match what you like and value.<br>• You feel like you can find other ways to reach your goals when things get in the way.<br>• You feel that the actions you take matter.<br><br>Some of the questions talk about **goals.**<br><br>• A goal is something you want to work towards, either now or in the future.<br>• Think about goals that are important to you, personally. These may be different than |

Copyright 2025, Academic Autism Spectrum Partnership in Research and Education (AASPIRE)

| | | |
|---|---|---|
| | | what other people feel is important.<br><br>The scale is **NOT** trying to measure disability.<br>• it is ok to use supports to do any of the things the questions ask about, as long as you **want** and value that support.<br>• It is trying to measure **things that may change over time**, for example, as someone gets older or gets different supports, therapies, training, accommodations, or resources.<br><br>Your goals may be different **in different parts of your life**. |
| **Additional Information** | | • Use this link for additional Information and examples to help you answer the questions, even if your answers may be different in different parts of your life. |
| **Response Options** | Disagree [slider] Agree | Strongly Disagree<br>Disagree<br>Neither Agree nor Disagree<br>Agree<br>Strongly Agree |
| **Item Changes** | I set my own goals. | I usually set my own goals (with as much support as I want). |
| | I have what it takes to reach my goals. | I think I have what I need to reach my goals. |

Copyright 2025, Academic Autism Spectrum Partnership in Research and Education (AASPIRE)

| | | |
|---|---|---|
| | I think of more than one way to solve a problem. | When I have a problem, I can usually think of more than one way to solve it. |
| | I consider many possibilities when I make plans for my future. | When I make plans for my future, I can usually think of many different possibilities. |
| | I know what I do best. | I know what I am good at and not so good at. |
| | I plan weekend activities I like to do. | I usually get to plan what to do with my free time. |
| | I keep trying even after I get something wrong. | I usually keep trying after I get something wrong. |
| | I think trying hard helps me get what I want. | In general, I believe that trying hard can help me reach my goals. |
| | I choose activities I want to do. | I usually choose to do activities I want to do, with as much support as I want. |
| | I work hard to reach my goals. | I usually work hard to reach my goals. |
| | I figure out ways to get around obstacles. | When something gets in my way, I can usually figure out ways to get around it. |
| | I am confident in my abilities. | I feel confident in my ability to do the things that are important to me, with whatever supports I want or need. |
| | My past experiences help me plan what I will do next. | The lessons I learned from my past experiences can often help me plan what I will do next. |
| | I think about each of my goals. | I often think about my goals. |
| | I make choices that are important to me. | I usually make choices that are important to me. |
| | I look for new experiences I think I will like. | I tend to look for new experiences I think I will like. |
| | I am able to focus to reach my goals. | I think that I will be able to reach my goals. |
| | I choose what my room looks like. | I can choose how to decorate my space within my home. |

Copyright 2025, Academic Autism Spectrum Partnership in Research and Education (AASPIRE)

| | | |
|---|---|---|
| | I take action when new opportunities come my way. | I usually take advantage of new opportunities when they come my way. |
| | I know my strengths. | I can recognize and understand my strengths. |
| | I come up with ways to reach my goals. | I can think of one or more ways to reach my goals. |
| **Hyperlinks** | • Possibilities<br>• Obstacles<br>• Opportunities | • I have what I need<br>• Goals<br>• Possibilities<br>• Opportunities<br>• Strengths |

Copyright 2025, Academic Autism Spectrum Partnership in Research and Education (AASPIRE)

SUPPLEMENTARY FILE
ARTICLE TITLE: Self- and Caregiver-Reported Choice-Making in Autistic Adults: Development and Validation of the AASPIRE–Choices and Decisions Scale

**Participants and Missing Data**

Thirty-one participants (3.6%) missed all items from both the SDI:AR-AASPIRE and AASPIRE–CDS instruments, and an additional nine participants (1.0%) missed all SDI:AR-AASPIRE items. Therefore, after excluding participants who did not complete any items, we included 830 participants in the analyses for the SDI:AR-AASPIRE (see Supplementary Table S1 for demographic characteristics for participants included in the SDI:AR-AASPIRE analyses). The proportion of missing responses per item ranged from 0.1% to 0.7% for SDI:AR-AASPIRE items (affecting 0–6 participants per item). Similar to the analyses in AASPIRE–CDS, we addressed missing data by generating 10 imputed datasets using a conditional multivariate normal distribution based on observed response patterns.

**Factor Structure of SDI:AR-AASPIRE**

To assess the internal structure of the SDI:AR-AASPIRE, we conducted a series of confirmatory factor analyses (CFA) within the SEM framework, testing two potential structural models. First, we examined a three-dimensional structure aligned with the original theoretical model of the SDI (Shogren et al., 2015). We then evaluated the factor correlations to determine whether to retain or reject the three-factor model. Second, we tested a unidimensional structure, given that prior research has also suggested the possibility of a single general factor of self-determination (Shogren et al., 2018). Because the factor correlations in the three-factor CFA model of SDI:AR-AASPIRE were very high (ranging from 0.93 to 1.00), we concluded that the factors were not sufficiently distinct and proceeded with a unidimensional model. The initial unidimensional model yielded RMSEA = 0.105, CFI = .789, TLI = .766, and SRMR = .067. We conducted a series of sensitivity checks to evaluate the impact of modification indices on standardized loadings and identified a revised model that included 23 error covariances and met the predefined fit criteria, yielding RMSEA = .055, CFI = .949, TLI = .935, and SRMR = .037.

After excluding the CR group, we re-estimated the models to evaluate the fit of the three-factor versus unidimensional structure. The factor correlations in the three-factor model were still high (ranging from 0.92 to 0.98). The global fit indices for the unidimensional model were: RMSEA = 0.106, CFI = .774, TLI = .749, and SRMR = .069. We conducted a series of sensitivity checks to evaluate the impact of modification indices on standardized loadings and identified a revised model that included 23 error covariances and met the predefined fit criteria, yielding RMSEA = .054, CFI = .948, TLI = .934, and SRMR = .039.

**CFA and Multi-Group CFA Results**

We conducted CFA separately by report type. The DR model met all predefined fit criteria. The DR-CS model met all criteria except the TLI criterion, which was only slightly below the predefined cutoff (TLI = 0.896 vs. 0.90), whereas the CR model did not meet multiple fit criteria (Supplementary Table S2). We therefore retained the DR-CS group in the primary report-type measurement invariance analyses but excluded the CR group. See Supplementary Table S2 for model fit results from the measurement invariance analyses using report type as the grouping variable. The full configural and metric invariance models met the predefined fit criteria. Although full scalar invariance was not supported, we achieved partial scalar invariance by freeing the intercept for one item, *Solutions*, resulting in fit indices that met the predefined criteria. The living-status-specific CFA models met the predefined fit criteria (Supplementary Table S2). For living status, configural and metric invariance were established, and partial scalar invariance was achieved by freeing the intercepts of two items when excluding the CR and four items when including the CR. We report the latent mean and variance differences by report type and living status in Supplementary Table S3.

We also conducted CFAs and multi-group CFAs with respect to communication fluency, subcohort, and gender, both including and excluding the CR group. Participant characteristics by communication fluency, subcohort, and gender are presented in Supplementary Table S4. CFA results and differences in latent means and variances are presented in Supplementary Tables S5 and S6, respectively.

**Reliabilities**

The SDI:AR-AASPIRE showed excellent internal consistency, with $\alpha = 0.92$ and $\omega = 0.92$ after excluding CR responses, and similarly high reliability when including CR responses ($\alpha = 0.93$; $\omega = 0.93$). Within the CR group, SDI:AR-AASPIRE demonstrated internal consistency with $\alpha = 0.91$ and $\omega = 0.91$. The test-retest reliability was 0.94.

SUPPLEMENTARY FILE

ARTICLE TITLE: Self- and Caregiver-Reported Choice-Making in Autistic Adults: Development and Validation of the AASPIRE–Choices and Decisions Scale

Supplementary Table S1.

*Demographic Characteristics by Grouping Variables Included in the Self-Determination Inventory:Adult Report-AASPIRE Analyses*

| Demographic Variable | Report Type | | | Living Status | |
|---|---|---|---|---|---|
| | DR (*n* = 586) | DR-CS (*n* = 183) | CR (*n* = 61) | I own/rent (*n* = 370) | Other (*n* = 457) |
| Mean Age (SD) [Range] | 33.25 (11.97) [18, 77] | 25.98 (9.48) [18, 75] | 25.23 (6.60) [18, 54] | 37.83 (12.62) [18, 77] | 25.61 (7.10) [18, 59] |
| | *n* (%) | *n* (%) | *n* (%) | *n* (%) | *n* (%) |
| Gender-Male | 183 (31.50%) | 89 (49.44%) | 51 (85.00%) | 81 (21.89%) | 242 (53.66%) |
| Gender-Female | 178 (30.64%) | 58 (32.22%) | 9 (15.00%) | 127 (34.32%) | 118 (26.16%) |
| Gender- Other | 220 (37.87%) | 33 (18.33%) | 0 (0%) | 162 (43.78%) | 91 (20.18%) |
| Race/Ethnicity | | | | | |
| White | 440 (75.99%) | 121 (66.85%) | 41 (68.33%) | 291 (79.51%) | 311 (68.50%) |
| Hispanic | 65 (11.23%) | 28 (15.47%) | 3 (5.00%) | 33 (9.02%) | 63 (13.88%) |
| Asian | 20 (3.45%) | 10 (5.52%) | 5 (8.33%) | 11 (3.01%) | 24 (5.29%) |
| Black | 13 (2.25%) | 11 (6.08%) | 6 (10.00%) | 4 (1.09%) | 26 (5.73%) |
| Other | 41 (7.08%) | 11 (6.08%) | 5 (8.33%) | 27 (7.38%) | 30 (6.61%) |
| Presence of ID | 59 (10.14%) | 71 (39.44%) | 50 (83.33%) | 30 (8.17%) | 150 (32.97%) |
| Report type | | | | | |
| DR | - | - | - | 346 (93.51%) | 239 (52.30%) |
| DR-CS | - | - | - | 24 (6.49%) | 158 (34.57%) |
| CR | - | - | - | 0 (0%) | 60 (13.13%) |
| Communication fluency | | | | | |
| Very basic ideas[a] | 14 (2.40%) | 11 (6.01%) | 30 (50.00%) | 2 (0.54%) | 53 (11.65%) |
| Simple ideas[b] | 10 (1.71%) | 23 (12.57%) | 14 (23.33%) | 8 (2.17%) | 39 (8.57%) |
| Full sentences, limited complexity[c] | 100 (17.12%) | 60 (32.79%) | 15 (25.00%) | 57 (15.45%) | 116 (25.49%) |
| Fluent[d] | 460 (78.77%) | 89 (48.63%) | 1 (1.67%) | 302 (81.84%) | 247 (54.29%) |
| Living Status | | | | | |
| I own/rent | 346 (59.15%) | 24 (13.19%) | 0 (0%) | - | - |
| Other | 239 (40.85%) | 158 (86.81%) | 60 (100.00%) | - | - |

*Notes*. Calculated based on the available data from 830 participants after excluding those who missed all items from the SDI:AR-AASPIRE. ID = Intellectual Disability. DR = Direct Report. DR-CS = Direct Report-Caregiver Support. CR = Caregiver Report. The numbers may not add up to 830 because there were missing data in the demographic information. [a] = I can only communicate very basic ideas or feelings; [b] = I can communicate a small number of simple ideas or feelings; [c] = I can communicate by putting together multiple sentences, but I have trouble discussing more complicated thoughts and ideas; [d] = I can communicate fluently.

SUPPLEMENTARY FILE
ARTICLE TITLE: Self- and Caregiver-Reported Choice-Making in Autistic Adults: Development and Validation of the AASPIRE–Choices and Decisions Scale

Supplementary Table S2.

*CFA Fit Indices for the SDI:AR-AASPIRE by Report Type and Living Status*

| Model | RMSEA | RMSEA 90% CI | CFI | TLI | SRMR | Tenable? | ΔCFI | AIC | Variables freed |
|---|---|---|---|---|---|---|---|---|---|
| Report Type (DR vs. DR-CS for the multigroup CFA) Excluding the CR for the Measurement Invariance Analyses | | | | | | | | | |
| DR | 0.054 | [0.048, 0.060] | 0.948 | 0.935 | 0.039 | N/A | N/A | N/A | |
| DR-CS | 0.069 | [0.057, 0.081] | 0.918 | 0.896 | 0.059 | N/A | N/A | N/A | |
| CR | 0.120 | [0.099, 0.141] | 0.764 | 0.602 | 0.095 | N/A | N/A | N/A | |
| Configural | 0.059 | [0.053, 0.064] | 0.940 | 0.924 | 0.050 | N/A | N/A | 38274.45 | - |
| Metric | 0.058 | [0.053, 0.064] | 0.937 | 0.925 | 0.071 | Yes | -0.003 | 38275.37 | - |
| Scalar | 0.062 | [0.057, 0.067] | 0.925 | 0.916 | 0.073 | No | -0.012 | 38341.88 | - |
| Scalar (P) | 0.061 | [0.056, 0.066] | 0.927 | 0.917 | 0.073 | Yes | -0.010 | 38329.90 | Solutions |
| Living Status (Own Arrangement vs. Other) Excluding the CR | | | | | | | | | |
| Own Arrangement | 0.055 | [0.047, 0.063] | 0.949 | 0.936 | 0.046 | N/A | N/A | N/A | |
| Other | 0.063 | [0.055, 0.070] | 0.929 | 0.910 | 0.049 | N/A | N/A | N/A | |
| Configural | 0.059 | [0.054, 0.064] | 0.939 | 0.923 | 0.048 | N/A | N/A | 38124.12 | - |
| Metric | 0.058 | [0.053, 0.064] | 0.937 | 0.925 | 0.058 | Yes | -0.002 | 38121.12 | - |
| Scalar | 0.063 | [0.058, 0.068] | 0.922 | 0.919 | 0.058 | No | -0.015 | 38208.16 | - |
| Scalar (P) | 0.061 | [0.056, 0.066] | 0.928 | 0.918 | 0.058 | Yes | -0.009 | 38171.75 | Decorate, Confidence |
| Living Status (Own Arrangement vs. Other) Including the CR | | | | | | | | | |
| Own Arrangement | 0.054 | [0.046, 0.062] | 0.950 | 0.937 | 0.045 | N/A | N/A | N/A | |
| Other | 0.062 | [0.055, 0.069] | 0.937 | 0.920 | 0.045 | N/A | N/A | N/A | |
| Configural | 0.059 | [0.053, 0.064] | 0.943 | 0.928 | 0.045 | N/A | N/A | 41498.13 | - |
| Metric | 0.060 | [0.055, 0.065] | 0.937 | 0.925 | 0.065 | Yes | -0.006 | 41526.64 | - |
| Scalar | 0.066 | [0.061, 0.071] | 0.919 | 0.909 | 0.067 | No | -0.018 | 41655.35 | - |
| Scalar (P) | 0.063 | [0.058, 0.067] | 0.928 | 0.918 | 0.066 | Yes | -0.009 | 41586.34 | Decorate, Confidence, Solutions, Reach Goals |

*Notes*. CFA = Confirmatory Factor Analysis; DR = Direct Report, DR-CS = Direct Report with Caregiver Support, CR = Caregiver Report. AIC = Akaike's Information Criterion. ΔCFI represents the change in CFI from the comparison model to the current model. Positive values indicate an increase in CFI, and negative values indicate a decrease in CFI. Partial scalar models were compared with the metric model. We considered models tenable when CFI decreased by no more than 0.01.

SUPPLEMENTARY FILE
ARTICLE TITLE: Self- and Caregiver-Reported Choice-Making in Autistic Adults: Development and Validation of the AASPIRE–Choices and Decisions Scale

Supplementary Table S3.

*Wald Test Results for Group Differences in SDI:AR-AASPIRE*

| Latent Variable | Groups | Latent Mean | χ2 | *p*-value | Latent Variances | χ2 | *p*-value |
|---|---|---|---|---|---|---|---|
| *Excluding the CR* | | | | | | | |
| Report Type | Direct Report | 4.21 | 3.65 | 0.06 | 0.21 | 0.18 | 0.67 |
| | Direct Report with Caregiver Support | 4.13 | | | 0.22 | | |
| Living Status | Own Arrangement | 4.20 | 1.14 | 0.28 | 0.23 | 0.66 | 0.42 |
| | Other | 4.16 | | | 0.21 | | |
| *Including the CR* | | | | | | | |
| Living Status | Own Arrangement | 4.17 | 13.17 | 0.0003 | 0.31 | 1.31 | 0.24 |
| | Other | 4.02 | | | 0.35 | | |

*Notes*. CR = Caregiver report.

SUPPLEMENTARY FILE
ARTICLE TITLE: Self- and Caregiver-Reported Choice-Making in Autistic Adults: Development and Validation of the AASPIRE–Choices and Decisions Scale

Supplementary Table S4.

*Demographic Characteristics by Grouping Variables Included in the SDI:AR-AASPIRE Analyses*

| Demographic Variable | *Cohort* | | | Communication Fluency | | Gender | | |
|---|---|---|---|---|---|---|---|---|
| | DDS (*n* = 258) | Healthcare (*n* = 292) | Community (*n* = 280) | Fluent (*n* = 550) | Non-fluent (*n* = 277) | Male (*n* = 323) | Female (*n* = 245) | Other (*n* = 253) |
| Mean Age (SD) [Range] | 26.68 (8.13) [18, 62] | 30.04 (11.79) [18, 77] | 36.15 (12.34) [19, 72] | 32.42 (11.99) [18, 75] | 28.36 (10.49) [18, 77] | 28.27 (10.04) [18, 75] | 34.54 (13.44) [18, 72] | 31.46 (10.88) [18, 77] |
| | *n* (%) | *n* (%) | *n* (%) | *n* (%) | *n* (%) | *n* (%) | *n* (%) | *n* (%) |
| Gender | | | | | | | | |
| Male | 148 (58.73%) | 128 (44.14%) | 47 (16.85%) | 180 (32.97%) | 142 (52.21%) | - | - | - |
| Female | 58 (23.02%) | 89 (30.69%) | 98 (35.13%) | 163 (29.85%) | 80 (29.41%) | - | - | - |
| Other | 46 (18.25%) | 73 (25.17%) | 134 (48.03%) | 203 (37.18%) | 50 (18.38%) | - | - | - |
| Race/Ethnicity | | | | | | | | |
| White | 161 (63.64%) | 211 (72.26%) | 230 (83.64%) | 428 (78.53%) | 172 (63.24%) | 222 (69.38%) | 177 (72.54%) | 199 (79.60%) |
| Hispanic | 47 (18.58%) | 28 (9.59%) | 21 (7.64%) | 51 (9.36%) | 44 (16.18%) | 44 (13.75%) | 25 (10.25%) | 25 (10.00%) |
| Asian | 15 (5.93%) | 13 (4.45%) | 7 (2.55%) | 19 (3.49%) | 16 (5.88%) | 16 (5.00%) | 13 (5.33%) | 6 (2.40%) |
| Black | 11 (4.35%) | 14 (4.79%) | 5 (1.82%) | 14 (2.57%) | 16 (5.88%) | 17 (5.31%) | 10 (4.10%) | 3 (1.20%) |
| Other | 19 (7.51%) | 26 (8.90%) | 12 (4.36%) | 33 (6.06%) | 24 (8.82%) | 21 (6.56%) | 19 (7.79%) | 17 (6.80%) |
| Presence of ID | 78 (30.71%) | 93 (31.85%) | 9 (3.26%) | 46 (8.42%) | 133 (48.72%) | 110 (34.38%) | 53 (21.63%) | 17 (6.77%) |
| Report type | | | | | | | | |

| | | | | | | | | |
|---|---|---|---|---|---|---|---|---|
| DR | 132 (51.16%) | 191 (65.41%) | 263 (93.93%) | 460 (83.64%) | 124 (44.77%) | 183 (56.66%) | 178 (72.65%) | 220 (86.96%) |
| DR-CS | 98 (37.98%) | 69 (23.63%) | 16 (5.71%) | 89 (16.18%) | 94 (33.94%) | 89 (27.55%) | 58 (23.67%) | 33 (13.04%) |
| CR | 28 (10.85%) | 32 (10.96%) | 1 (0.36%) | 1 (0.18%) | 59 (21.30%) | 51 (15.79%) | 9 (3.67%) | 0 (0.00%) |
| Communication fluency | | | | | | | | |
| Very basic ideas[a] | 20 (7.81%) | 32 (10.96%) | 3 (1.08%) | - | - | 39 (12.11%) | 11 (4.53%) | 5 (1.98%) |
| Simple ideas[b] | 24 (9.38%) | 21 (7.19%) | 2 (0.72%) | - | - | 30 (9.32%) | 9 (3.70%) | 7 (2.77%) |
| Full sentences, limited complexity[c] | 77 (30.08%) | 63 (21.58%) | 35 (12.54%) | - | - | 73 (22.67%) | 60 (24.69%) | 38 (15.02%) |
| Fluent[d] | 135 (52.73%) | 176 (60.27%) | 239 (85.66%) | - | - | 180 (55.90%) | 163 (67.08%) | 203 (80.24%) |
| Living Status | | | | | | | | |
| I own/rent | 54 (21.09%) | 114 (39.04%) | 202 (72.40%) | 302 (55.01%) | 67 (24.36%) | 81 (25.08%) | 127 (51.84%) | 162 (64.03%) |
| Other | 202 (78.91%) | 178 (60.96%) | 77 (27.60%) | 247 (44.99%) | 208 (75.64%) | 242 (74.92%) | 118 (48.16%) | 91 (35.97%) |

*Notes*. Calculated based on the available data from 830 participants after excluding those who missed all items from the SDI–AASPIRE. ID = Intellectual Disability. DR = Direct Report. DR-CS = Direct Report-Caregiver Support. CR = Caregiver Report. The numbers may not add up to 830 because there were missing data in the demographic information. [a] = I can only communicate very basic ideas or feelings; [b] = I can communicate a small number of simple ideas or feelings; [c] = I can communicate by putting together multiple sentences, but I have trouble discussing more complicated thoughts and ideas; [d] = I can communicate fluently. For communication fluency, we dichotomized responses into two groups: non-fluent communication (i.e., individuals who can communicate basic needs, short phrases, or sentences but have difficulty expressing complex ideas) and fluent communication. We dichotomized the communication fluency variable because over 65% of participants were classified as fluent, and alternative categorization approaches would have led to model non-convergence in measurement invariance testing.

SUPPLEMENTARY FILE
ARTICLE TITLE: Self- and Caregiver-Reported Choice-Making in Autistic Adults: Development and Validation of the AASPIRE–Choices and Decisions Scale

Supplementary Table S5.

*CFA Fit Indices for the SDI:AR-AASPIRE by Communication Fluency, Subcohort, and Gender*

| Model | RMSEA | RMSEA 90% CI | CFI | TLI | SRMR | Tenable? | ΔCFI | AIC | Variables freed |
|---|---|---|---|---|---|---|---|---|---|
| Communication Fluency (Fluent Communication vs. Non-fluent Communication) Excluding the CR | | | | | | | | | |
| Non-fluent Communication | 0.067 | [0.056, 0.078] | 0.914 | 0.891 | 0.054 | N/A | N/A | N/A | |
| Fluent Communication | 0.058 | [0.052, 0.064] | 0.942 | 0.927 | 0.042 | N/A | N/A | N/A | |
| Configural | 0.060 | [0.055, 0.066] | 0.935 | 0.917 | 0.048 | N/A | N/A | 38115.40 | - |
| Metric | 0.059 | [0.054, 0.064] | 0.934 | 0.921 | 0.058 | Yes | -0.001 | 38097.98 | - |
| Scalar | 0.059 | [0.054, 0.064] | 0.930 | 0.921 | 0.059 | Yes | -0.004 | 38110.28 | |
| Communication Fluency (Fluent Communication vs. Non-fluent Communication) Including the CR | | | | | | | | | |
| Non-fluent Communication | 0.060 | [0.051, 0.070] | 0.934 | 0.917 | 0.046 | N/A | N/A | N/A | |
| Fluent Communication | 0.057 | [0.051, 0.063] | 0.943 | 0.928 | 0.041 | N/A | N/A | N/A | |
| Configural | 0.058 | [0.053, 0.063] | 0.940 | 0.924 | 0.044 | N/A | N/A | 41412.23 | - |
| Metric | 0.058 | [0.053, 0.063] | 0.936 | 0.923 | 0.066 | Yes | -0.004 | 41425.66 | - |
| Scalar | 0.062 | [0.057, 0.066] | 0.925 | 0.915 | 0.066 | No | -0.011 | 41491.25 | |
| Scalar (P) | 0.061 | [0.056, 0.066] | 0.927 | 0.917 | 0.066 | Yes | -0.009 | 41476.04 | Reach goals |
| Cohort (DDS vs. Healthcare vs. Community) Excluding the CR | | | | | | | | | |
| DDS | 0.065 | [0.054, 0.075] | 0.935 | 0.918 | 0.049 | N/A | N/A | N/A | |
| Healthcare | 0.061 | [0.051, 0.071] | 0.936 | 0.919 | 0.050 | N/A | N/A | N/A | |
| Community | 0.059 | [0.049, 0.068] | 0.935 | 0.918 | 0.050 | N/A | N/A | N/A | |
| Configural | 0.061 | [0.056, 0.067] | 0.936 | 0.919 | 0.050 | N/A | N/A | 38236.12 | |
| Metric | 0.059 | [0.054, 0.065] | 0.934 | 0.923 | 0.064 | Yes | -0.002 | 38206.76 | |
| Scalar | 0.065 | [0.060, 0.070] | 0.916 | 0.908 | 0.064 | No | -0.018 | 38301.65 | |
| Scalar (P) | 0.062 | [0.057, 0.067] | 0.924 | 0.916 | 0.064 | Yes | -0.010 | 38248.12 | Solutions, Lessons, Reach Goals, Confidence |
| Cohort (DDS vs. Healthcare vs. Community) Including the CR | | | | | | | | | |
| DDS | 0.059 | [0.050, 0.069] | 0.947 | 0.933 | 0.044 | N/A | N/A | N/A | |

| | | | | | | | | | |
|---|---|---|---|---|---|---|---|---|---|
| Healthcare | 0.063 | [0.054, 0.072] | 0.937 | 0.921 | 0.047 | N/A | N/A | N/A | |
| Community | 0.059 | [0.049, 0.068] | 0.935 | 0.918 | 0.050 | N/A | N/A | N/A | |
| Configural | 0.060 | [0.055, 0.066] | 0.940 | 0.924 | 0.047 | N/A | N/A | 41730.50 | - |
| Metric | 0.060 | [0.054, 0.065] | 0.937 | 0.926 | 0.066 | Yes | -0.003 | 41716.74 | - |
| Scalar | 0.066 | [0.061, 0.071] | 0.917 | 0.910 | 0.066 | No | -0.020 | 41838.27 | - |
| Scalar (P) | 0.062 | [0.057, 0.067] | 0.927 | 0.919 | 0.066 | Yes | -0.010 | 41767.71 | Solutions, Lessons, Reach Goals, Confidence, Think Goals |
| Gender (Male vs. Female vs. Other) Excluding the CR | | | | | | | | | |
| Male | 0.058 | [0.047, 0.068] | 0.938 | 0.922 | 0.048 | N/A | N/A | N/A | |
| Female | 0.066 | [0.056, 0.075] | 0.927 | 0.908 | 0.049 | N/A | N/A | N/A | |
| Other | 0.063 | [0.053, 0.072] | 0.932 | 0.914 | 0.052 | N/A | N/A | N/A | |
| Configural | 0.062 | [0.056, 0.068] | 0.932 | 0.914 | 0.050 | N/A | N/A | 37889.40 | |
| Metric | 0.061 | [0.055, 0.066] | 0.931 | 0.919 | 0.067 | Yes | -0.001 | 37861.05 | |
| Scalar | 0.064 | [0.059, 0.070] | 0.916 | 0.908 | 0.067 | No | -0.015 | 37927.07 | |
| Scalar (P) | 0.063 | [0.057, 0.068] | 0.921 | 0.913 | 0.067 | Yes | -0.010 | 37895.45 | Believe, Work Hard, Decorate |
| Gender (Male vs. Female vs. Other) Including the CR | | | | | | | | | |
| Male | 0.067 | [0.059, 0.075] | 0.933 | 0.916 | 0.044 | N/A | N/A | N/A | |
| Female | 0.055 | [0.045, 0.066] | 0.946 | 0.932 | 0.044 | N/A | N/A | N/A | |
| Other | 0.062 | [0.052, 0.072] | 0.933 | 0.915 | 0.052 | N/A | N/A | N/A | |
| Configural | 0.062 | [0.057, 0.068] | 0.937 | 0.920 | 0.047 | N/A | N/A | 41310.46 | - |
| Metric | 0.061 | [0.056, 0.066] | 0.935 | 0.923 | 0.066 | Yes | -0.002 | 41289.39 | - |
| Scalar | 0.067 | [0.062, 0.072] | 0.916 | 0.908 | 0.066 | No | -0.019 | 41405.40 | - |
| Scalar (P) | 0.064 | [0.059, 0.069] | 0.925 | 0.916 | 0.066 | Yes | -0.010 | 41344.64 | Believe, Work Hard, Decorate, Needs Met, Activities, Solutions, Problem Solve |

*Notes*. CFA = Confirmatory Factor Analysis; DDS = Developmental Disability Services. For communication fluency, we dichotomized responses into two groups: non-fluent communication (i.e., individuals who can communicate basic needs, short phrases, or sentences but have difficulty expressing complex ideas) and fluent communication. We dichotomized the communication fluency variable because over 65% of participants were classified as fluent, and alternative categorization approaches would have led to model non-convergence in measurement invariance testing. AIC = Akaike's Information Criterion. ΔCFI represents the change in CFI from the comparison model to the current model. Positive values indicate an increase in CFI, and negative values indicate a decrease in CFI. Partial scalar models were compared with the metric model. We considered models tenable when CFI decreased by no more than 0.01.

SUPPLEMENTARY FILE
ARTICLE TITLE: Self- and Caregiver-Reported Choice-Making in Autistic Adults: Development and Validation of the AASPIRE–Choices and Decisions Scale

Supplementary Table S6.

*Wald Test Results for Group Differences in SDI:AR-AASPIRE by Communication Fluency, Subcohort, and Gender*

| Latent Variable | Groups | Latent Mean | χ2 | *p*-value | Latent Variances | χ2 | *p*-value |
|---|---|---|---|---|---|---|---|
| *Excluding the CR* | | | | | | | |
| Communication Fluency | Fluent Communication | 4.25 | 33.59 | < 0.0001 | 0.20 | 0.02 | 0.88 |
| | Non-fluent Communication | 4.02 | | | 0.20 | | |
| Cohort | DDS | 4.24 | 5.07 | 0.08 | 0.22 | 0.90 | 0.64 |
| | Healthcare | 4.13 | | | 0.22 | | |
| | Community | 4.20 | | | 0.20 | | |
| Gender | Male | 4.18 | 7.32 | 0.03 | 0.21 | 0.64 | 0.73 |
| | Female | 4.24 | | | 0.19 | | |
| | Other | 4.13 | | | 0.21 | | |
| *Including the CR* | | | | | | | |
| Communication Fluency | Fluent Communication | 4.23 | 71.05 | < 0.0001 | 0.25 | 2.87 | 0.09 |
| | Non-fluent Communication | 3.84 | | | 0.31 | | |
| Cohort | DDS | 4.12 | 9.19 | 0.01 | 0.35 | 6.37 | 0.04 |
| | Healthcare | 4.00 | | | 0.36 | | |
| | Community | 4.15 | | | 0.26 | | |
| Gender | Male | 4.08 | 0.31 | 0.86 | 0.35 | 2.00 | 0.37 |
| | Female | 4.10 | | | 0.32 | | |
| | Other | 4.09 | | | 0.29 | | |

*Notes*. DDS = Developmental Disability Services. For communication fluency, we dichotomized responses into two groups: non-fluent communication (i.e., individuals who can communicate basic needs, short phrases, or sentences but have difficulty expressing complex ideas) and fluent communication. We dichotomized the communication fluency variable because over 65% of participants were classified as fluent, and alternative categorization approaches would have led to model non-convergence in measurement invariance testing.

#### Supplementary Materials B

##### Selection of Choices Construct and Instrument Development

The CBPR-nested Delphi process in the project's 1st phase consisted of 4 cycles, each of which included a survey of study team members and Delphi panelists, a shared decision-making meeting with the study team, and creation of materials for the next round of Delphi surveys (details in Nicolaidis et al, 2025). In the first two rounds of Delphi surveys, we reduced an initial list of 60 potential outcomes to 30, asked Delphi panelists to help define each construct, and solicited recommendations for existing instruments to measure high-priority outcomes. During Round 3, we asked Delphi panelists to review the Basic Psychological Needs Satisfaction Scale (La Guardia et al, 2000) and the SDI:AR, and answer a series of questions about which they preferred and if and how each instrument would need to be adapted for autistic adults and our focal construct. While Delphi panelists generally preferred the SDI:AR and felt that it only needed minor adaptations, several noted that neither instrument did a good job at capturing what was most important to them about self-determination, namely whether they even had the *opportunity* to make meaningful choices. In Round 4, we compared the SDI:AR to the Choice subscale of NCI–ACS. Continued disagreement among panelists led us to realize that we would need to include two measures of self-determination, one such as the SDI:AR to capture internal psychological aspects, and another such as the NCI-ACS to capture opportunities to make choices.

During Phase 2, we reviewed existing measures related to the opportunity to make choices (e.g., the NCI–ACS Choice subscale; Restrictiveness Evaluation Measure for Youth; Rauktis et al., 2009), but team members raised concerns about their applicability to autistic adults with diverse support needs. Several raised concerns about the NCI–ACS's applicability to autistic adults who do not receive formal disability services (e.g., items that presume a case manager or other formal provider). Team members also reviewed the Restrictiveness Evaluation Measure for Youth, which assesses environmental restrictiveness in child care and residential service settings, including limits on activities (e.g., clothing), movement (e.g., where a person can go within the care setting), and social interactions (e.g., contact with friends), as well as constraints embedded in treatment (e.g., treatment intensity) and restrictions related to independent living (e.g., finances; Rauktis et al., 2009), as not well-suited to our aims because it assumes residential care structures and emphasizes restrictions rather than meaningful opportunities to make choices and decisions. As such, we chose to make minor adaptations to the SDI:AR and create a new scale, intended as a complement to the SDI:AR-AASPIRE, to measure the opportunity to make choices and decisions.

We initially created a new instrument (with direct-report and caregiver-report versions) that we called "The Freedom to Make Choices Scale." It included 13 items focused on different life domains (e.g., living situation, friendships, job or career) and listed examples of the types of choices or decisions one might make within each domain. However, cognitive interviews with autistic adults and caregivers revealed several problems. First, some participants had trouble responding because they were able to make some of the choices or decisions listed as examples within a given domain, but not others. For example, they may be able to choose how to decorate their room, but they can't choose where to live. As such, we significantly restructured the instrument so that each item only focused on a specific choice or decision. We included one item from each of the 13 domains, but we created items that asked about choices and decisions with a range of magnitude and risk. Initial items focus on small choices that do not carry a lot of risk (e.g. "I usually get to choose what I wear") with gradually growing magnitude or risk as the instrument progresses (e.g., "I usually get to decide when and where to go when I leave my home." to "I usually get to make my own decisions about my job or career.") .

Additionally, a caregiver felt "attacked" because she felt like the instrument was implying that she *should* be giving her loved one the opportunity to make choices and decisions that they were not *able* to make. For example, she felt that framing the construct as "freedom" to make choices implied it is something everyone should have. We renamed the measure as AASPIRE-CDS and removed any language referring to "freedom," instead phrasing questions to ask about whether the person "gets to" make the specific choice or decision. We also added text to the preface emphasizing that participants may not be able or allowed to make all their own choices or decisions, and that more freedom to make choices or decisions is not necessarily better in all circumstances. Cognitive interviews using the revised measure were much more positive, requiring only trivial additional revisions.

Across this process, 23 participants (17 autistic adults and six caregivers or supporters, including one autistic adult and one supporter who participated together as a dyad) reviewed the choices and decisions measure during cognitive interviews. Twelve autistic adults and two caregivers reviewed the initial Freedom to Make Choices Scale. Four autistic adults, three caregivers, and the dyad reviewed the restructured version as it was further refined into the AASPIRE-CDS.

SUPPLEMENTARY FILE
ARTICLE TITLE: Self- and Caregiver-Reported Choice-Making in Autistic Adults: Development and Validation of the AASPIRE–Choices and Decisions Scale

#### Supplementary Materials C

#### Information Presented in the Clickable Links

##### Examples of Things that May Limit Your Ability to Make Your Own Choices or Decisions

The following are some examples of things that may limit your ability to make your own choices or decisions:

- Not being able to understand the choices, or the risks and benefits related to those
- choices.
- People not telling you that there is a decision to be made or what all the choices are.
- People not letting you have a say in the decision.
- People ignoring what you say or choose.
- People making you do something you really don't want to do.
- People not letting you do something you can and want to do.
- People not understanding your likes, dislikes, priorities, choices, or decisions.
- Having to follow rules, restrictions, or limitations that you don't believe are reasonable.
- Feeling like you have less choices than other people because of discrimination, abuse, or
- extreme pressure from others.
- Not having the information, supports, accommodations, time, or relationships you need
- to make a decision or do an activity.
- Not being allowed to make choices that others feel are risky or unwise.

##### More Information On Why Making Your Own Choices Or Decisions Doesn't Mean That

##### You Can Do Whatever You Want.

Making your own choices or decisions doesn't mean you can do whatever you want. It can still include:

- Following rules or restrictions, as long as you feel they are reasonable.
- Making a choice you don't necessarily like, for example because you feel it's the right
- thing to do.
- Being limited by laws and rules that apply to everyone (not just people with disabilities).

##### More Information About What It Means To Make Your Own Choices Or Decisions

##### With Support From Others.

Making your own choices or decisions doesn't mean you have to do it alone. It can still include:

- Listening to other people's advice or recommendations, as long as you get to choose
- whether or not to follow the advice.
- Getting help from other people to make decisions or do activities, as long as you want
- the help.

SUPPLEMENTARY FILE
ARTICLE TITLE: Self- and Caregiver-Reported Choice-Making in Autistic Adults: Development and Validation of the AASPIRE–Choices and Decisions Scale

**Supplementary Materials D**

**Caregiver Information Sources and Confidence Ratings for the AASPIRE-CDS and SDI:AR-AASPIRE, and Support Provided for Direct Reports**

Supplementary Table S7.

*How Caregivers Knew the Information Used to Complete the AASPIRE-CDS and SDI:AR-AASPIRE*

| How do you know this information? (Check all that apply.) | AASPIRE-CDS, n (%) (n = 63) | SDI:AR-AASPIRE, n (%) (n = 55) |
|---|---|---|
| They told me (in whatever mode they communicated) | 13 (20.6%) | 13 (23.6%) |
| I have observed the action, behavior, or event | 58 (92.1%) | 52 (94.5%) |
| I can interpret their behaviors and/or vocalizations | 38 (60.3%) | 35 (63.6%) |
| Someone else told me | 7 (11.1%) | 8 (14.5%) |
| I feel like I can guess because I have known them for a long time | 33 (52.4%) | 36 (65.5%) |
| I feel like I can guess because I spend a lot of time with them | 34 (54.0%) | 30 (54.5%) |
| Other | 2 (3.2%) | 0 (0.0%) |

*Note*. Percentages were calculated based on the number of caregiver respondents who completed the item for each instrument. Respondents could select more than one option.

SUPPLEMENTARY FILE
ARTICLE TITLE: Self- and Caregiver-Reported Choice-Making in Autistic Adults: Development and Validation of the AASPIRE–Choices and Decisions Scale

Supplementary Table S8.

*Confidence Ratings for Choices and Self-Determination (0–10 Scale)*

| On a scale of 0 to 10, how confident are you about your response? | *n* (%) | |
|---|---|---|
| | *AASPIRE-Choices and Decisions* | *Self-Determination Inventory:Adult Report-AASPIRE* |
| 0 | 0 (0%) | 1 (1.8%) |
| 1 | 0 (0%) | 0 (0%) |
| 2 | 0 (0%) | 0 (0%) |
| 3 | 0 (0%) | 0 (0%) |
| 4 | 0 (0%) | 0 (0%) |
| 5 | 1 (1.8%) | 3 (5.5%) |
| 6 | 2 (3.6%) | 3 (5.5%) |
| 7 | 4 (7.3%) | 5 (9.1%) |
| 8 | 7 (12.7%) | 10 (18.2%) |
| 9 | 12 (21.8%) | 16 (29.1%) |
| 10 | 29 (52.7%) | 17 (30.9%) |

*Notes.* Percentage calculated based on the *n* of participants in Caregiver Report. Caregivers reported greater confidence in their AASPIRE-CDS ratings ($M$ = 9.07, $SD$ = 1.26) than in their SDI:AR-AASPIRE ratings ($M$ = 8.42, $SD$ = 1.76), p = .003.

SUPPLEMENTARY FILE
ARTICLE TITLE: Self- and Caregiver-Reported Choice-Making in Autistic Adults: Development and Validation of the AASPIRE–Choices and Decisions Scale

Supplementary Table S9.

*Type of Support Provided in Direct-Report with Caregiver Support*

| Type of Support Provided | *n* (%) | |
|---|---|---|
| | *AASPIRE-Choices and Decisions* | *Self-Determination Inventory:Adult Report-AASPIRE* |
| Someone helped me use a computer, smartphone, or other device (for example, they clicked on the answers I chose). | 41 (22.4%) | 40 (21.9%) |
| Someone read the survey to me. | 47 (25.7%) | 51 (27.9%) |
| Someone helped me understand what the questions or answers options meant. | 54 (29.5%) | 58 (31.7%) |
| Someone helped me decide how to answer the questions on the survey. | 19 (10.4%) | 24 (13.1%) |
| I got some other type of help to answer the questions. | 1 (0.5%) | 4 (2.2%) |
| Someone answered the questions for me, without my input. | 6 (3.3%) | 6 (3.3%) |

*Notes.* % calculated based on the *n* of direct-report with caregiver support. Respondents could select more than one type of support. Among the six participants who indicated that someone answered questions without their input across the two instruments, three endorsed at least one additional support option for the AASPIRE-CDS and four endorsed at least one additional support option for the SDI:AR-AASPIRE. We therefore treated the remaining three participants for the AASPIRE-CDS and two participants for the SDI:AR-AASPIRE, who did not endorse any other support option, as having had the instrument completed without their input. We repeated the relevant analyses after excluding these participants and obtained results consistent with the primary analyses.

### Supplementary Materials E

#### Secondary Measurement Invariance Analyses of AASPIRE-CDS With Regard to Subcohort, Gender, and Communication Fluency

##### Rationale for Conducting the Secondary Analysis in Regards to Subcohort, Gender, and Communication Fluency

We considered autistic participants' gender because the CR group included a substantially higher proportion of males (83.58%) than females (16.42%). We examined the subcohorts because findings from the broader project suggested systematic differences in experiences across subcohort types. We included communication fluency because it may be associated with individuals' opportunities to express preferences and advocate for themselves (Sturrock et al., 2022).

SUPPLEMENTARY FILE
ARTICLE TITLE: Self- and Caregiver-Reported Choice-Making in Autistic Adults: Development and Validation of the AASPIRE–Choices and Decisions Scale

Supplementary Table S10.

*Demographic Characteristics by Subcohort, Communication Fluency, and Gender Included in the AASPIRE-CDS Analyses*

| Demographic Variable | Cohort | | | Communication Fluency | | Gender | | |
|---|---|---|---|---|---|---|---|---|
| | DDS (*n* = 260) | Healthcare (*n* = 299) | Community (*n* = 280) | Fluent (*n* = 551) | Non-fluent (*n* = 285) | Male (*n* = 328) | Female (*n* = 247) | Other (*n* = 253) |
| Mean Age (SD) [Range] | 26.66 (8.11) [18, 62] | 29.84 (11.73) [18, 77] | 36.15 (12.34) [19, 72] | 32.42 (11.98) [18, 75] | 28.17 (10.43) [18, 77] | 28.18 (10.00) [18, 62] | 34.42 (13.45) [18, 77] | 31.46 (10.88) [18, 77] |
| | *n* (%) | *n* (%) | *n* (%) | *n* (%) | *n* (%) | *n* (%) | *n* (%) | *n* (%) |
| Gender | | | | | | | | |
| Male | 149 (58.89%) | 132 (44.59%) | 47 (16.85%) | 180 (32.97%) | 147 (52.69%) | - | - | - |
| Female | 58 (22.92%) | 91 (30.74%) | 98 (35.13%) | 163 (29.85%) | 82 (29.39%) | - | - | - |
| Other | 46 (18.18%) | 73 (24.66%) | 134 (48.03%) | 203 (37.18%) | 50 (17.92%) | - | - | - |
| Race/Ethnicity | | | | | | | | |
| White | 163 (63.92%) | 214 (71.81%) | 230 (83.64%) | 428 (78.53%) | 177 (63.21%) | 225 (69.23%) | 178 (72.36%) | 199 (79.60%) |
| Hispanic | 47 (18.43%) | 29 (9.73%) | 21 (7.64%) | 51 (9.36%) | 45 (16.07%) | 45 (13.85%) | 25 (10.16%) | 25 (10.00%) |
| Asian | 15 (5.88%) | 13 (4.36%) | 7 (2.55%) | 19 (3.49%) | 16 (5.71%) | 16 (4.92%) | 13 (5.28%) | 6 (2.40%) |
| Black | 11 (4.31%) | 15 (5.03%) | 5 (1.82%) | 14 (2.57%) | 17 (6.07%) | 17 (5.23%) | 11 (4.47%) | 3 (1.20%) |
| Other | 19 (7.45%) | 27 (9.06%) | 12 (4.36%) | 33 (6.06%) | 25 (8.93%) | 22 (6.77%) | 19 (7.72%) | 17 (6.80%) |
| Presence of ID | 79 (30.86%) | 98 (32.89%) | 9 (3.26%) | 46 (8.42%) | 139 (49.47%) | 114 (35.08%) | 55 (22.27%) | 17 (6.77%) |
| Report type | | | | | | | | |
| DR | 132 (50.77%) | 192 (64.21%) | 263 (93.93%) | 461 (83.67%) | 124 (43.51%) | 183 (55.79%) | 178 (72.06%) | 220 (86.96%) |
| DR-CS | 98 (37.69%) | 69 (23.08%) | 16 (5.71%) | 89 (16.15%) | 94 (32.98%) | 89 (27.13%) | 58 (23.48%) | 33 (13.04%) |

| | | | | | | | | |
|---|---|---|---|---|---|---|---|---|
| CR | 30 (11.54%) | 38 (12.71%) | 1 (0.36%) | 1 (0.18%) | 67 (23.51%) | 56 (17.07%) | 11 (4.45%) | 0 (0.00%) |
| Communication fluency | | | | | | | | |
| Very basic ideas[a] | 20 (7.75%) | 36 (12.04%) | 3 (1.08%) | - | - | 42 (12.84%) | 12 (4.90%) | 5 (1.98%) |
| Simple ideas[b] | 25 (9.69%) | 22 (7.36%) | 2 (0.72%) | - | - | 32 (9.79%) | 9 (3.67%) | 7 (2.77%) |
| Full sentences, limited complexity[c] | 78 (30.23%) | 64 (21.40%) | 35 (12.54%) | - | - | 73 (22.32%) | 61 (24.90%) | 38 (15.02%) |
| Fluent[d] | 135 (52.33%) | 177 (59.20%) | 239 (85.66%) | - | - | 180 (55.05%) | 163 (66.53%) | 203 (80.24%) |
| Living Status | | | | | | | | |
| I own/rent | 54 (20.93%) | 114 (38.26%) | 202 (72.40%) | 302 (55.01%) | 67 (23.67%) | 81 (24.70%) | 127 (51.42%) | 162 (64.03%) |
| Other | 204 (79.07%) | 184 (61.74%) | 77 (27.60%) | 247 (44.99%) | 216 (76.33%) | 247 (75.30%) | 120 (48.58%) | 91 (35.97%) |

*Notes*. ID = Intellectual Disability. DR = Direct Report. DR-CS = Direct Report-Caregiver Support. CR = Caregiver Report. The numbers may not add up to 839 because there were missing data in the demographic information. [a] = I can only communicate very basic ideas or feelings; [b] = I can communicate a small number of simple ideas or feelings; [c] = I can communicate by putting together multiple sentences, but I have trouble discussing more complicated thoughts and ideas; [d] = I can communicate fluently. For communication fluency, we dichotomized responses into two groups: non-fluent communication (i.e., individuals who can communicate basic needs, short phrases, or sentences but have difficulty expressing complex ideas) and fluent communication. We dichotomized the communication fluency variable because over 65% of participants were classified as fluent, and alternative categorization approaches would have led to model non-convergence in measurement invariance testing.

SUPPLEMENTARY FILE
ARTICLE TITLE: Self- and Caregiver-Reported Choice-Making in Autistic Adults: Development and Validation of the AASPIRE–Choices and Decisions Scale

Supplementary Table S11.

*CFA Fit Indices for the AASPIRE-CDS by Communication Fluency, Subcohort, and Gender*

| Model | RMSEA | RMSEA 90% CI | CFI | TLI | SRMR | Tenable? | ΔCFI | AIC | Variables freed |
|---|---|---|---|---|---|---|---|---|---|
| Communication Fluency (Fluent Communication vs. Non-fluent Communication) Excluding the CR | | | | | | | | | |
| Non-fluent Communication | 0.065 | [0.045, 0.084] | 0.947 | 0.919 | 0.050 | N/A | N/A | N/A | |
| Fluent Communication | 0.064 | [0.053, 0.075] | 0.959 | 0.938 | 0.035 | N/A | N/A | N/A | |
| Configural | 0.064 | [0.055, 0.074] | 0.957 | 0.934 | 0.043 | N/A | N/A | 22902.42 | |
| Metric | 0.062 | [0.053, 0.072] | 0.954 | 0.937 | 0.059 | Yes | -0.003 | 22899.62 | |
| Scalar | 0.061 | [0.052, 0.070] | 0.951 | 0.940 | 0.059 | Yes | -0.003 | 22897.79 | |
| Communication Fluency (Fluent Communication vs. Non-fluent Communication) Including the CR | | | | | | | | | |
| Non-fluent Communication | 0.072 | [0.056, 0.088] | 0.955 | 0.931 | 0.045 | N/A | N/A | N/A | |
| Fluent Communication | 0.064 | [0.053, 0.075] | 0.959 | 0.938 | 0.035 | N/A | N/A | N/A | |
| Configural | 0.067 | [0.058, 0.076] | 0.958 | 0.936 | 0.040 | N/A | N/A | 25590.78 | |
| Metric | 0.066 | [0.058, 0.075] | 0.953 | 0.936 | 0.059 | Yes | -0.005 | 25599.41 | |
| Scalar | 0.066 | [0.058, 0.075] | 0.947 | 0.935 | 0.057 | Yes | -0.006 | 25622.97 | |
| Cohort (DDS vs. Healthcare vs. Community) Excluding the CR | | | | | | | | | |
| DDS | 0.077 | [0.059, 0.095] | 0.941 | 0.910 | 0.048 | N/A | N/A | N/A | |
| Healthcare | 0.074 | [0.057, 0.091] | 0.954 | 0.929 | 0.042 | N/A | N/A | N/A | |
| Community | 0.075 | [0.060, 0.092] | 0.934 | 0.899 | 0.052 | N/A | N/A | N/A | |
| Configural | 0.075 | [0.066, 0.085] | 0.944 | 0.914 | 0.048 | N/A | N/A | 22932.07 | |
| Metric | 0.075 | [0.066, 0.084] | 0.936 | 0.915 | 0.072 | Yes | -0.008 | 22939.61 | |
| Scalar | 0.080 | [0.072, 0.088] | 0.916 | 0.903 | 0.073 | No | -0.020 | 22991.93 | |
| Scalar (P) | 0.076 | [0.068, 0.084] | 0.926 | 0.912 | 0.073 | Yes | -0.010 | 22957.96 | Free Time, Touch |
| Cohort (DDS vs. Healthcare vs. Community) Including the CR | | | | | | | | | |
| DDS | 0.062 | [0.045, 0.080] | 0.964 | 0.946 | 0.041 | N/A | N/A | N/A | |

| | | | | | | | | | |
|---|---|---|---|---|---|---|---|---|---|
| Healthcare | 0.066 | [0.050, 0.082] | 0.972 | 0.957 | 0.035 | N/A | N/A | N/A | |
| Community | 0.075 | [0.059, 0.091] | 0.936 | 0.901 | 0.052 | N/A | N/A | N/A | |
| Configural | 0.068 | [0.059, 0.077] | 0.961 | 0.940 | 0.043 | N/A | N/A | 25846.99 | |
| Metric | 0.070 | [0.061, 0.078] | 0.952 | 0.937 | 0.073 | Yes | -0.009 | 25865.29 | |
| Scalar | 0.075 | [0.067, 0.083] | 0.937 | 0.926 | 0.076 | No | -0.015 | 25919.90 | - |
| Scalar (P) | 0.071 | [0.063, 0.079] | 0.945 | 0.935 | 0.075 | Yes | -0.007 | 25880.96 | Free Time, Touch |
| Gender (Male vs. Female vs. Other) Excluding the CR | | | | | | | | | |
| Male | 0.067 | [0.049, 0.085] | 0.953 | 0.928 | 0.043 | N/A | N/A | N/A | |
| Female | 0.062 | [0.045, 0.079] | 0.961 | 0.941 | 0.041 | N/A | N/A | N/A | |
| Other | 0.087 | [0.071, 0.103] | 0.932 | 0.896 | 0.050 | N/A | N/A | N/A | |
| Configural | 0.073 | [0.063, 0.082] | 0.948 | 0.921 | 0.045 | N/A | N/A | 22863.41 | |
| Metric | 0.072 | [0.063, 0.081] | 0.941 | 0.922 | 0.072 | Yes | -0.007 | 22869.17 | |
| Scalar | 0.073 | [0.064, 0.081] | 0.932 | 0.920 | 0.072 | Yes | -0.009 | 22881.45 | |
| Gender (Male vs. Female vs. Other) Including the CR | | | | | | | | | |
| Male | 0.067 | [0.052, 0.081] | 0.970 | 0.954 | 0.034 | N/A | N/A | N/A | |
| Female | 0.073 | [0.056, 0.090] | 0.948 | 0.920 | 0.044 | N/A | N/A | N/A | |
| Other | 0.086 | [0.070, 0.103] | 0.933 | 0.897 | 0.050 | N/A | N/A | N/A | |
| Configural | 0.075 | [0.066, 0.084] | 0.954 | 0.930 | 0.043 | N/A | N/A | 25751.86 | |
| Metric | 0.074 | [0.065, 0.082] | 0.948 | 0.932 | 0.070 | Yes | -0.006 | 25757.19 | |
| Scalar | 0.075 | [0.067, 0.083] | 0.939 | 0.929 | 0.071 | Yes | -0.009 | 25782.11 | |

*Notes*. DDS = Developmental Disability Services. CR = Caregiver Report. For communication fluency, we dichotomized responses into two groups: non-fluent communication (i.e., individuals who can communicate basic needs, short phrases, or sentences but have difficulty expressing complex ideas) and fluent communication. We dichotomized the communication fluency variable because over 65% of participants were classified as fluent, and alternative categorization approaches would have led to model non-convergence in measurement invariance testing. AIC = Akaike's Information Criterion. ΔCFI represents the change in CFI from the comparison model to the current model. Positive values indicate an increase in CFI, and negative values indicate a decrease in CFI. Partial scalar models were compared with the metric model. We considered models tenable when CFI decreased by no more than 0.01.

SUPPLEMENTARY FILE
ARTICLE TITLE: Self- and Caregiver-Reported Choice-Making in Autistic Adults: Development and Validation of the AASPIRE–Choices and Decisions Scale

Supplementary Table S12.

*Wald Test Results for Group Differences in AASPIRE-CDS by Communication Fluency, Subcohort, and Gender*

| Latent Variable | Groups | Latent Mean | χ2 | *p*-value | Latent Variances | χ2 | *p*-value |
|---|---|---|---|---|---|---|---|
| *Excluding the CR* | | | | | | | |
| Communication Fluency | Fluent Communication | 4.27 | 56.07 | < 0.0001 | 0.39 | 0.31 | 0.58 |
| | Non-fluent Communication | 3.83 | | | 0.42 | | |
| Cohort | DDS | 4.02 | 33.68 | < 0.0001 | 0.43 | 16.59 | 0.0002 |
| | Healthcare | 4.07 | | | 0.50 | | |
| | Community | 4.33 | | | 0.28 | | |
| Gender | Male | 4.20 | 2.12 | 0.35 | 0.38 | 2.76 | 0.25 |
| | Female | 4.11 | | | 0.44 | | |
| | Other | 4.16 | | | 0.48 | | |
| *Including the CR* | | | | | | | |
| Communication Fluency | Fluent Communication | 4.27 | 125.11 | < 0.0001 | 0.40 | 18.26 | < 0.0001 |
| | Non-fluent Communication | 3.54 | | | 0.72 | | |
| Cohort | DDS | 3.89 | 76.59 | < 0.0001 | 0.58 | 45.60 | < 0.0001 |
| | Healthcare | 3.84 | | | 0.83 | | |
| | Community | 4.33 | | | 0.29 | | |
| Gender | Male | 3.84 | 24.56 | < 0.0001 | 0.84 | 19.70 | 0.0001 |
| | Female | 4.14 | | | 0.46 | | |
| | Other | 4.16 | | | 0.51 | | |

*Notes*. DDS = Developmental Disability Services. CR = Caregiver Report. For communication fluency, we dichotomized responses into two groups: non-fluent communication (i.e., individuals who can communicate basic needs, short phrases, or sentences but have difficulty expressing complex ideas) and fluent communication. We dichotomized the communication fluency variable because over 65% of participants were classified as fluent, and alternative categorization approaches would have led to model non-convergence in measurement invariance testing.

SUPPLEMENTARY FILE
ARTICLE TITLE: Self- and Caregiver-Reported Choice-Making in Autistic Adults: Development and Validation of the AASPIRE–Choices and Decisions Scale

#### Supplementary Materials F

#### Assumptions for Pearson Correlation Analyses

Examination of normality assumptions, based on visual inspection and Shapiro–Wilk tests, indicated that the independent living needs and communication fluency variables (coded as a four-level variable) showed non-normal distributions, and AASPIRE–CDS, personal care needs, SDI:AR-AASPIRE, and validity indicators for SDI:AR-AASPIRE (i.e., quality of life, overall health, and employment satisfaction) met the assumptions of normality. However, we used Pearson's correlation due to its general robustness to violations of normality in large samples (Field, 2018), and because results using Spearman's rho differed minimally from those obtained with Pearson's correlation.

#### Supplementary Materials G

#### Identification of Factor Structures of AASPIRE-CDS Including and Excluding the Caregiver Report

In the EFA on the split-half sample after excluding the CR, we extracted two factors, explaining 52.16% of the total variance, with an average KMO value of 0.92 and Factors 1 and 2 yielding eigenvalues of 5.63 and 1.15, respectively and a clear elbow in the scree plot supporting the unidimensional structure (Supplementary Table S13). In the EFA re-specifying one factor, all 13 items loaded onto Factor 1, explaining 43.33% of the total variance, and Factor 1 had an eigenvalue of 5.63 (Supplementary Table S14). The CFA on the other half showed adequate model fit after adding 14 error covariances (RMSEA = 0.071, CFI = 0.954, TLI = 0.930, SRMR = 0.039; standardized factor loadings ranged from 0.51 to 0.73, see Supplementary Table S15 for standardized loading for each item).

SUPPLEMENTARY FILE
ARTICLE TITLE: Self- and Caregiver-Reported Choice-Making in Autistic Adults: Development and Validation of the AASPIRE–Choices and Decisions Scale

Supplementary Table S13.

*Results of the Initial Exploratory Factor Analysis of AASPIRE-CDS*

| Items | Including the CR Group | | Excluding the CR Group | |
|---|---|---|---|---|
| | Factor 1 | Factor 2 | Factor 1 | Factor 2 |
| Wear | | 0.72 | | 0.68 |
| Decorate | | 0.84 | | 0.82 |
| Free Time | | 0.73 | | 0.70 |
| Friends | 0.54 | 0.48 | 0.41 | 0.52 |
| Room | 0.41 | 0.62 | 0.31 | 0.66 |
| Communicate | 0.59 | | 0.66 | |
| Outside Home | 0.48 | 0.55 | 0.50 | 0.56 |
| Touch | 0.40 | 0.44 | | 0.48 |
| Relationships | 0.73 | 0.31 | 0.62 | 0.32 |
| Money | 0.56 | 0.46 | 0.57 | 0.37 |
| Health | 0.80 | | 0.76 | |
| Job | 0.82 | | 0.79 | |
| Political | 0.76 | 0.31 | 0.68 | |

*Note. CR = Caregiver-report.*

SUPPLEMENTARY FILE
ARTICLE TITLE: Self- and Caregiver-Reported Choice-Making in Autistic Adults: Development and Validation of the AASPIRE–Choices and Decisions Scale

Supplementary Table S14.

*Results of the Re-specified One-Factor Exploratory Factor Analysis of the AASPIRE-CDS*

| Items | Including the CR Group | Excluding the CR Group |
|---|---|---|
| Wear | 0.68 | 0.62 |
| Decorate | 0.70 | 0.68 |
| Free Time | 0.64 | 0.63 |
| Friends | 0.72 | 0.66 |
| Room | 0.72 | 0.68 |
| Communicate | 0.65 | 0.60 |
| Outside Home | 0.78 | 0.75 |
| Touch | 0.59 | 0.49 |
| Relationships | 0.75 | 0.67 |
| Money | 0.73 | 0.67 |
| Health | 0.74 | 0.67 |
| Job | 0.75 | 0.71 |
| Political | 0.77 | 0.70 |

*Note. CR = Caregiver-report.*

SUPPLEMENTARY FILE
ARTICLE TITLE: Self- and Caregiver-Reported Choice-Making in Autistic Adults: Development and Validation of the AASPIRE–Choices and Decisions Scale

Supplementary Table S15.

*CFA Standardized Loadings of the Final Model of the AASPIRE-CDS from the Confirmatory Factor Analysis*

| | Standardized Loading | |
|---|---|---|
| Items | Including the CR group | Excluding the CR group |
| Job | 0.69 | 0.61 |
| Health | 0.76 | 0.69 |
| Politics | 0.78 | 0.72 |
| Relationships | 0.73 | 0.70 |
| Money | 0.75 | 0.73 |
| Communicate | 0.63 | 0.56 |
| Outside Home | 0.66 | 0.62 |
| Friends | 0.75 | 0.72 |
| Decorate | 0.55 | 0.55 |
| Room | 0.65 | 0.64 |
| Free Time | 0.67 | 0.71 |
| Wear | 0.56 | 0.51 |
| Touch | 0.60 | 0.65 |

*Note. CR = Caregiver-report.*

SUPPLEMENTARY FILE
ARTICLE TITLE: Self- and Caregiver-Reported Choice-Making in Autistic Adults: Development and Validation of the AASPIRE–Choices and Decisions Scale

Supplementary Figure S1.

*Scree Plot of Eigenvalues for the AASPIRE-CDS Excluding the CR*

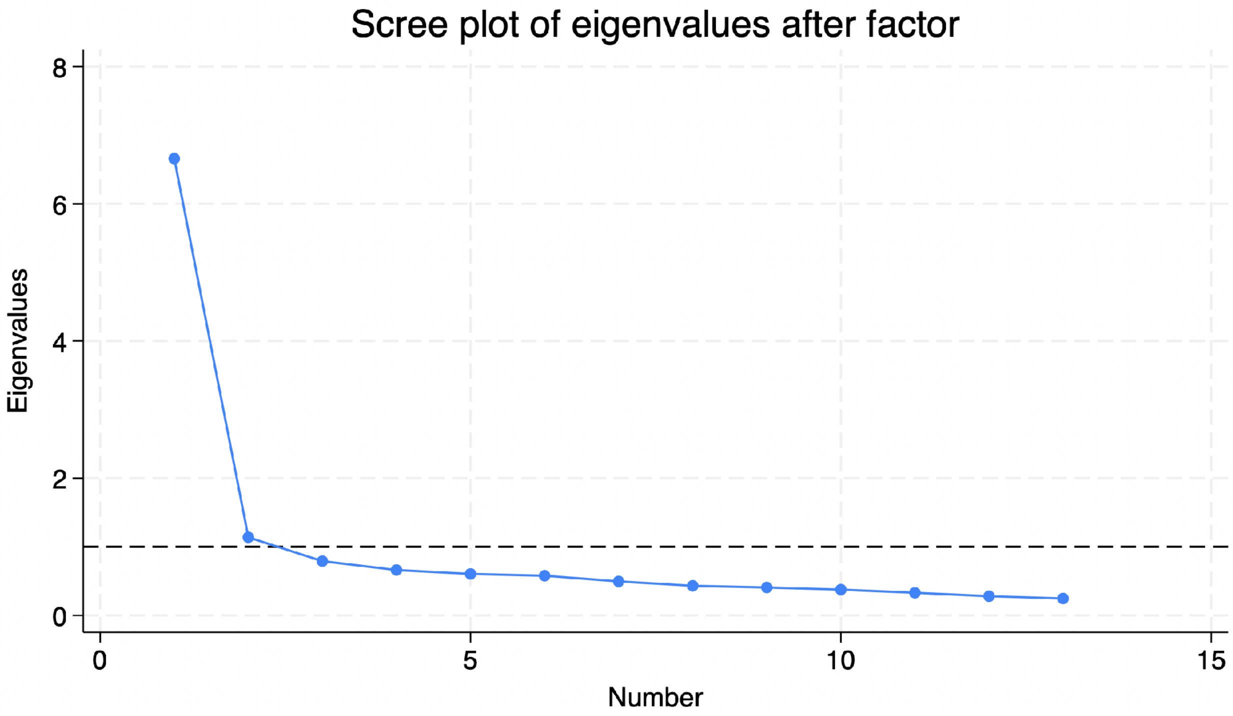

SUPPLEMENTARY FILE
ARTICLE TITLE: Self- and Caregiver-Reported Choice-Making in Autistic Adults: Development and Validation of the AASPIRE–Choices and Decisions Scale
